# The *TH01* microsatellite and *INS* VNTR are strongly associated with type 2 diabetes and fasting insulin secretion

**DOI:** 10.1101/2022.09.23.22280080

**Authors:** Jaime Berumen, Lorena Orozco, Héctor Gallardo-Rincón, Eligia Juárez-Torres, Elizabeth Barrera, Miguel Cruz-López, Rosa Elba Benuto, Espiridión Ramos-Martinez, Melissa Marin-Madina, Anabel Alvarado-Silva, Adán Valladares-Salgado, José de Jesús Peralta-Romero, Humberto García-Ortiz, Luis Alberto Martinez-Juarez, Alejandra Montoya, Diego-Abelardo Alvarez-Hernández, Jesús Alegre-Diaz, Pablo Kuri-Morales, Roberto Tapia-Conyer

## Abstract

A variable number of tandem repeats (VNTR) located in the insulin gene (*INS*) control region may be involved in the development of type 2 diabetes (T2D). The *TH01* microsatellite is located close to *INS* and has previously been suggested to be involved its regulation. Therefore, this observational study investigated whether the *TH01* microsatellite and *INS* VNTR, as assessed via the surrogate marker single nucleotide polymorphism (SNP) rs689, are associated with T2D in the Mexican population. Logistic regression models were used to calculate the risk conferred by *TH01* and *INS* VNTR loci for T2D development. *TH01* alleles 6, 8, 9 and 9.3 and allele A of rs689 were independently associated with T2D; differences were found between age at T2D diagnosis and sex. Larger alleles of *TH01* (≥8 repeats) conferred an increased risk for T2D in males when compared with smaller alleles (≤7 repeats) (odds ratio, ≥1.46; 95% confidence interval, 1.1–1.95). In females, larger alleles conferred a 1.5-fold higher risk for T2D when diagnosed at ≥46 years whereas they conferred protection when diagnosed at ≤45 years. Both *TH01* and SNP rs689 were associated with T2D in the same groups; the association remained significant for both loci in multivariate models. The median fasting plasma insulin concentration was significantly higher in patients with T2D versus controls, and in those diagnosed at ≤45 versus ≥46 years. *TH01* larger alleles or the A allele of rs689 may potentiate insulin synthesis in males, but not females, without T2D, a process that is disabled in those with T2D.

## Introduction

Type 2 diabetes (T2D) is associated with insulin resistance in peripheral tissues and a deficiency in insulin production by the pancreas [1, 2], which may be due to poor gene expression [3], mRNA maturation [4], or abnormal insulin secretion [5] by pancreatic β-cells. Single nucleotide polymorphisms (SNPs) located in genes involved in insulin maturation and production, such as *WFS1* [6] and *IGF2BP2* [7], or the regulation of insulin gene (*INS*) expression, such as *TCF7L2,* confer an increased risk for T2D [8]. The SNP rs149483638, which confers a lower risk for T2D, is located in the *INS-IGF2* gene located near the *INS* gene [9]. These data appear to suggest that the *INS* control region could be influenced directly or indirectly by proteins encoded either by these or other genes, or by nearby polymorphisms. In the *INS* control region, which begins upstream of the *INS* gene transcription start site, there is a variation in the number of tandem repeats (VNTR) polymorphism at 390 base pairs (bp) from the start of *INS* transcription (**Figure 1**) that is associated with type 1 diabetes [10–12]. The *INS* VNTR consists of a sequence of 14–15 nucleotides (5′-ACAGGGGTGTGGGG-3′) repeated in tandem [13] and has been shown to participate in the regulation of the *INS* in healthy individuals [14–16]. *INS* VNTR alleles are categorized according to the number of repeats (class I, 26–63 repetitions, average 570 bp; class II, 64–140 repetitions, average 1200 bp; class III, 141–209 repetitions, average 2200 bp). While class I *INS* VNTR alleles are considered to confer a risk of developing type 1 diabetes, class III alleles are generally associated with dominant protection [13]. Unlike class III alleles, class I alleles have been associated with increased insulin production [17, 18].

**Figure 1.**
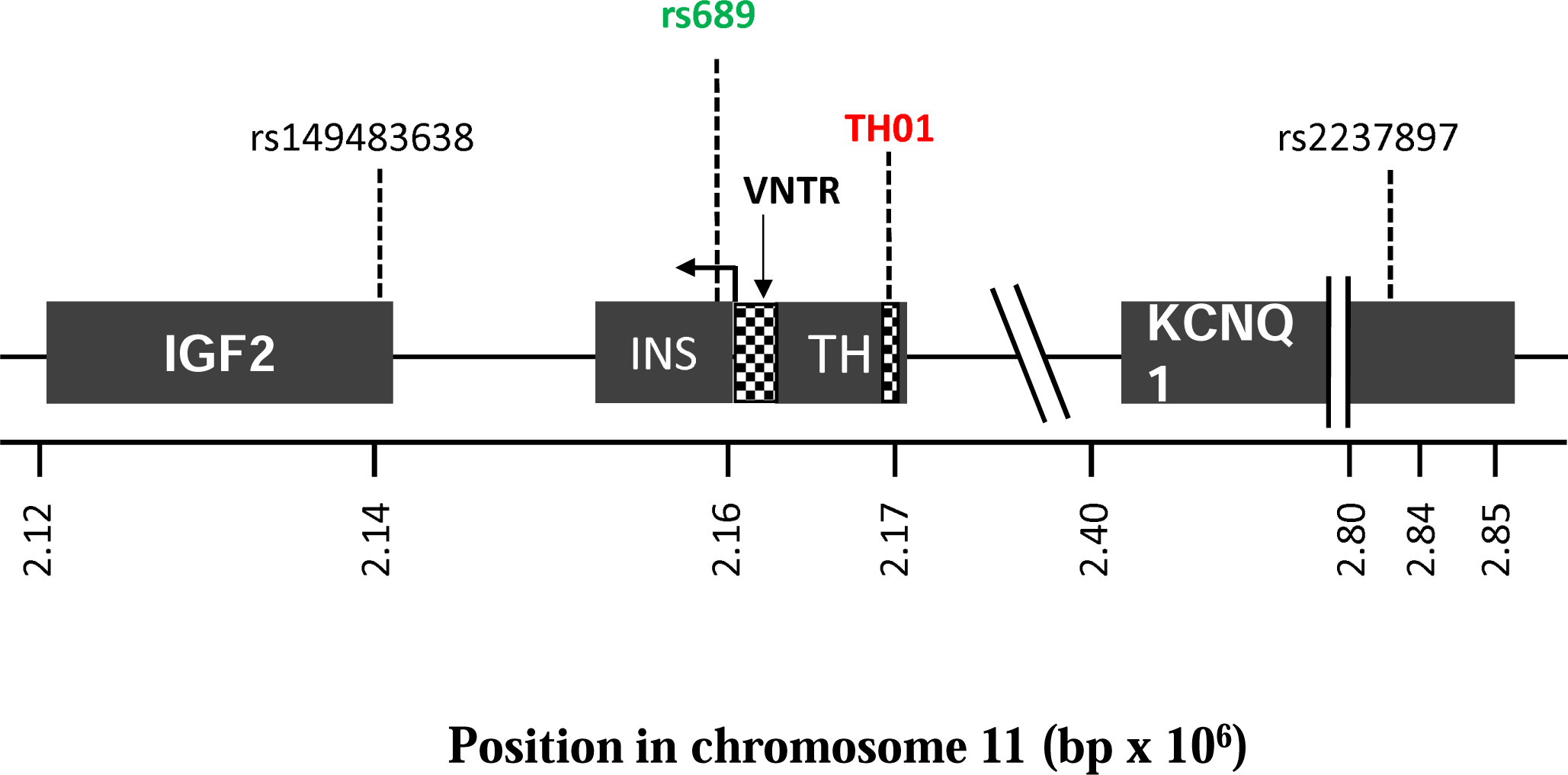
Genomic map of region 11p15.5. The figure shows the locations of genes *IGF2*, *INS*, *TH*, and *KCNQ1* and the markers SNP rs689, VNTR *INS*, and *TH01* microsatellite. The positions are based on the Human genome version GRCh38. The arrow indicates the direction and site of initiation of gene transcription. bp, base pairs; SNP, single nucleotide polymorphism; VNTR, variable number of tandem repeats.

Although the *INS* VNTR has been associated with T2D [19–22], the results remain largely controversial. Moreover, because most studies were conducted in European populations, there is little evidence in Latin Americans. There could be different associations between *INS* VNTR and T2D in the latter population, as polymorphisms in *SLC16A11*, *INS-IGF2*, and *HNF1A* genes have previously been reported to have strong associations with T2D in Mexicans, but not in Caucasians [23, 24].

The *INS* VNTR is complex; therefore, a surrogate marker, the SNP rs689, has been frequently used [22]. This marker is found within intron 1 of *INS* and is in complete linkage disequilibrium with *INS* VNTR in White populations (allele A is associated with class III alleles; allele T, with class I alleles). In addition, the tyrosine hydroxylase (*TH)01* microsatellite, a tetranucleotide (AATG) repeated 3–11 times in tandem, is also located relatively close to *INS* (approximately 9,800 bp). It is located within the *TH* gene that is in partial linkage imbalance with the *INS* VNTR in White [10, 25] and Japanese [26] populations. In addition, *in vitro* experiments have demonstrated that *TH01* has enhancer functions, i.e., it can regulate gene expression at a distance, a common attribute for microsatellites [27], and could potentially influence *INS* expression [28].

Interestingly, although *TH01* is associated with obesity [29], hypertension [30, 31], coronary heart disease [32], metabolic syndrome and high triglyceride levels [33, 34], its association with T2D has not been explored. Therefore, this study was conducted to investigate whether the *TH01* microsatellite and *INS* VNTR (through SNP rs689) are associated with T2D and the concentration of fasting plasma insulin in the Mexican population. The main study conducted was a case–control study that used univariate (ULR) and multivariate (MLR) logistic regression models to calculate the risk (OR, odd’s ratio) conferred of each loci for T2D in a Mexican population; a clinical replica case–control study and a cross-sectional study were also included.

## Results

### Participants and demographic characteristics

The main case–control study, replica case–control study, and cross-sectional study included 1986, 1188, and 1172 participants, respectively (**Table S1**); 53.3%, 60.6%, and 73.5% of participants were female, respectively. In both case–control studies, nearly 50% of the participants were T2D cases (i.e., 10.5% [main case–control study] and 3.2% [replica case– control study] of participants were diagnosed during recruitment, while the remaining cases had been previously diagnosed). In contrast, in the cross-sectional study, the prevalence of T2D was 5.4%; the majority (92/104) were diagnosed during recruitment, while only 12 cases had been previously diagnosed. At the time of enrollment, the mean ± standard deviation (SD) age of controls vs cases was similar across the studies (main, 59.2 ± 11.3 vs 55.6 ± 11.7 years; replica, 54.1 ± 10.1 vs 57.1 ± 9.5 years; cross-sectional, 49.1 ± 10.8 vs 49.2 ± 11.4 years), and the mean age at the time of T2D diagnosis was also similar across the studies (ranging from 44.8 to 49.2 years). The mean duration of disease was 9.5 ± 8.7 years in the main study, 12.3 ± 7.7 years in the replica case–control study, and approximately 0 years in the cross-sectional study (**Table S1**).

### Distribution of TH01 alleles stratified by sex and age at T2D diagnosis

*TH01* alleles were only assessed in the main study (*n =* 3972 chromosomes). The *TH01* allele frequency between cases and controls is summarized in **Table 1**. Though we identified 9 of the 11 alleles reported for the *TH01* microsatellite [35], only five (6, 7, 8, 9 and 9.3) had a frequency ≥5% in cases or controls. Whereas allele 6 was observed more frequently in controls than in cases (36.6% vs 30.2%, *P <* 0.0001), alleles 8 (5.2% vs 3.9%, *P <* 0.05) and 9.3 (21.0% vs 17.3%, *P* < 0.01) were more frequent among cases than controls. The difference between controls vs cases for allele 6 was greater in males (38.3% vs 29.4%; *P* < 0.0001) than in females (35.1% vs 30.9%; *P* < 0.05). Differences between cases vs controls for alleles 8 and 9.3 were only significant among males (5.8% vs 3.2%; *P* < 0.01 and 21.4% vs 17%; *P* < 0.05, respectively).

**Table 1.**
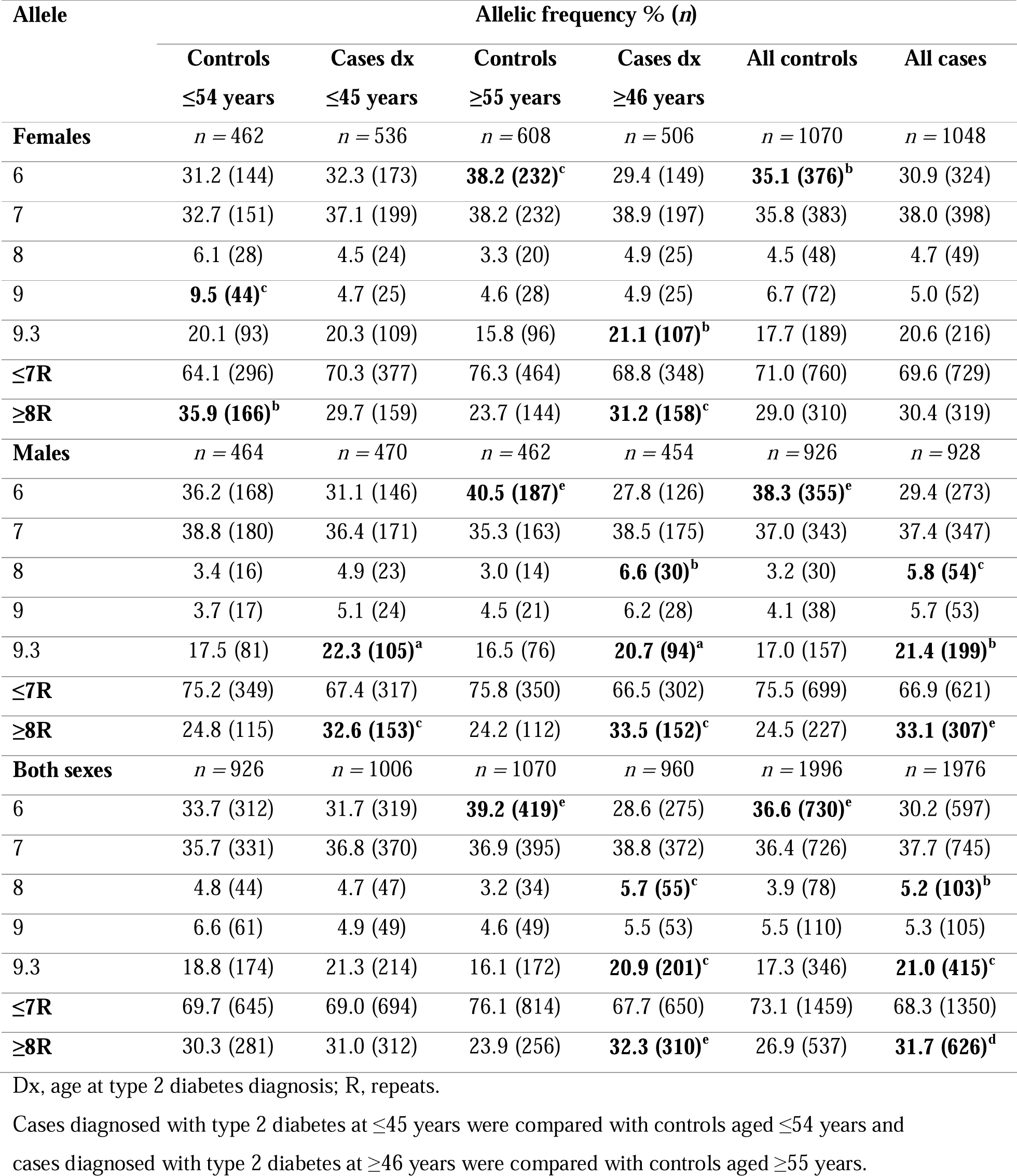
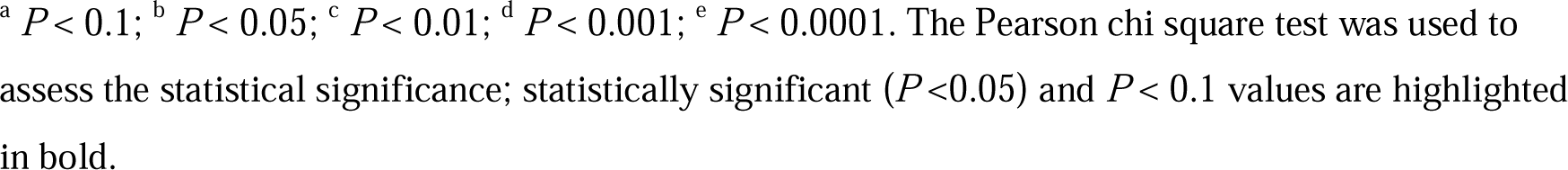
Comparison of frequency of *TH01* alleles with a minor allele frequency ≥0.05 between controls and cases (*n =* 3972).

| Allele | Allelic frequency % ($n$) | | | | | |
| --- | --- | --- | --- | --- | --- | --- |
| | Controls<br>$\leq 54$ years | Cases dx<br>$\leq 45$ years | Controls<br>$\geq 55$ years | Cases dx<br>$\geq 46$ years | All controls | All cases |
| **Females** | $n = 462$ | $n = 536$ | $n = 608$ | $n = 506$ | $n = 1070$ | $n = 1048$ |
| 6 | 31.2 (144) | 32.3 (173) | **38.2 (232)<sup>c</sup>** | 29.4 (149) | **35.1 (376)<sup>b</sup>** | 30.9 (324) |
| 7 | 32.7 (151) | 37.1 (199) | 38.2 (232) | 38.9 (197) | 35.8 (383) | 38.0 (398) |
| 8 | 6.1 (28) | 4.5 (24) | 3.3 (20) | 4.9 (25) | 4.5 (48) | 4.7 (49) |
| 9 | **9.5 (44)<sup>c</sup>** | 4.7 (25) | 4.6 (28) | 4.9 (25) | 6.7 (72) | 5.0 (52) |
| 9.3 | 20.1 (93) | 20.3 (109) | 15.8 (96) | **21.1 (107)<sup>b</sup>** | 17.7 (189) | 20.6 (216) |
| **$\leq 7R$** | 64.1 (296) | 70.3 (377) | 76.3 (464) | 68.8 (348) | 71.0 (760) | 69.6 (729) |
| **$\geq 8R$** | **35.9 (166)<sup>b</sup>** | 29.7 (159) | 23.7 (144) | **31.2 (158)<sup>c</sup>** | 29.0 (310) | 30.4 (319) |
| **Males** | $n = 464$ | $n = 470$ | $n = 462$ | $n = 454$ | $n = 926$ | $n = 928$ |
| 6 | 36.2 (168) | 31.1 (146) | **40.5 (187)<sup>e</sup>** | 27.8 (126) | **38.3 (355)<sup>e</sup>** | 29.4 (273) |
| 7 | 38.8 (180) | 36.4 (171) | 35.3 (163) | 38.5 (175) | 37.0 (343) | 37.4 (347) |
| 8 | 3.4 (16) | 4.9 (23) | 3.0 (14) | **6.6 (30)<sup>b</sup>** | 3.2 (30) | **5.8 (54)<sup>c</sup>** |
| 9 | 3.7 (17) | 5.1 (24) | 4.5 (21) | 6.2 (28) | 4.1 (38) | 5.7 (53) |
| 9.3 | 17.5 (81) | **22.3 (105)<sup>a</sup>** | 16.5 (76) | **20.7 (94)<sup>a</sup>** | 17.0 (157) | **21.4 (199)<sup>b</sup>** |
| **$\leq 7R$** | 75.2 (349) | 67.4 (317) | 75.8 (350) | 66.5 (302) | 75.5 (699) | 66.9 (621) |
| **$\geq 8R$** | 24.8 (115) | **32.6 (153)<sup>c</sup>** | 24.2 (112) | **33.5 (152)<sup>c</sup>** | 24.5 (227) | **33.1 (307)<sup>e</sup>** |
| **Both sexes** | $n = 926$ | $n = 1006$ | $n = 1070$ | $n = 960$ | $n = 1996$ | $n = 1976$ |
| 6 | 33.7 (312) | 31.7 (319) | **39.2 (419)<sup>e</sup>** | 28.6 (275) | **36.6 (730)<sup>e</sup>** | 30.2 (597) |
| 7 | 35.7 (331) | 36.8 (370) | 36.9 (395) | 38.8 (372) | 36.4 (726) | 37.7 (745) |
| 8 | 4.8 (44) | 4.7 (47) | 3.2 (34) | **5.7 (55)<sup>c</sup>** | 3.9 (78) | **5.2 (103)<sup>b</sup>** |
| 9 | 6.6 (61) | 4.9 (49) | 4.6 (49) | 5.5 (53) | 5.5 (110) | 5.3 (105) |
| 9.3 | 18.8 (174) | 21.3 (214) | 16.1 (172) | **20.9 (201)<sup>c</sup>** | 17.3 (346) | **21.0 (415)<sup>c</sup>** |
| **$\leq 7R$** | 69.7 (645) | 69.0 (694) | 76.1 (814) | 67.7 (650) | 73.1 (1459) | 68.3 (1350) |
| **$\geq 8R$** | 30.3 (281) | 31.0 (312) | 23.9 (256) | **32.3 (310)<sup>e</sup>** | 26.9 (537) | **31.7 (626)<sup>d</sup>** |

Dx, age at type 2 diabetes diagnosis; R, repeats.
Cases diagnosed with type 2 diabetes at $\leq 45$ years were compared with controls aged $\leq 54$ years and cases diagnosed with type 2 diabetes at $\geq 46$ years were compared with controls aged $\geq 55$ years.
<sup>a</sup> $P < 0.1$ ; <sup>b</sup> $P < 0.05$ ; <sup>c</sup> $P < 0.01$ ; <sup>d</sup> $P < 0.001$ ; <sup>e</sup> $P < 0.0001$ . The Pearson chi square test was used to assess the statistical significance; statistically significant ( $P < 0.05$ ) and $P < 0.1$ values are highlighted in bold.

When stratified by sex and age at diagnosis, the difference in the frequency of allele 6 increased in the older controls, irrespective of sex. In the older group, the difference in allele 8 frequency was significantly increased in male cases vs controls. Moreover, the difference of allele 9.3 frequency between controls and cases was significant in older females and in males regardless of age at T2D diagnosis. Similarly, allele 9 was more frequent in younger female controls vs cases. The large (L) alleles, those with ≥8 repeats (R; alleles 8, 9, 9.3, and 11) were predominant in older male and female cases, as well as in younger male cases, while the small (S) alleles, those with ≤7R (alleles 7, 6, 5, 4, and 3) predominated in the younger female cases.

### Association of TH01 with T2D using regression models

The association of *TH01* alleles 6, 8, 9, and 9.3 (*n =* 3972 chromosomes) with T2D is presented in **Table S2**. Allele 6 had a significantly protective effect towards T2D diagnosed at ≥46 years, with a 32.0% reduced risk in females and 44.0% in males, and allele 9 for T2D in females diagnosed at ≤45 years (54.0%). The opposite effect was observed for allele 8, which conferred a 2.26-times greater risk for T2D diagnosed at ≥46 years in males, and allele 9.3, which conferred a 1.43-times increased risk in females diagnosed at ≥46 years and a 1.34-times increased risk in males diagnosed at either age cutoff. The alleles with ≥8R conferred a similarly increased risk for T2D diagnosed at ≥46 years in both males (1.57 times) and females (1.46 times). Moreover, while they conferred a 1.46-times increased risk for T2D diagnosed at ≤45 years in males, they offered protection (25% reduction) in females diagnosed at ≤45 years.

The association of *TH01* genotypes (*n =* 1986 chromosomes) with T2D stratified by age at T2D diagnosis and sex is highlighted in **Table 2**. The associations of T2D with genotype were of greater magnitude than observed with alleles; the homozygous L/L genotypes conferred a much higher risk than heterozygous L/S genotypes. The risk in males increased from an odds ratio (OR) of 1.48 (95% confidence interval [CI], 1.13–1.95; *P =* 0.005) when heterozygous to 2.26 (95% CI, 1.41–3.64; *P =* 0.0007) when homozygous, indicating an additive effect with the number of alleles with ≥8R in the genotype.

**Table 2.**
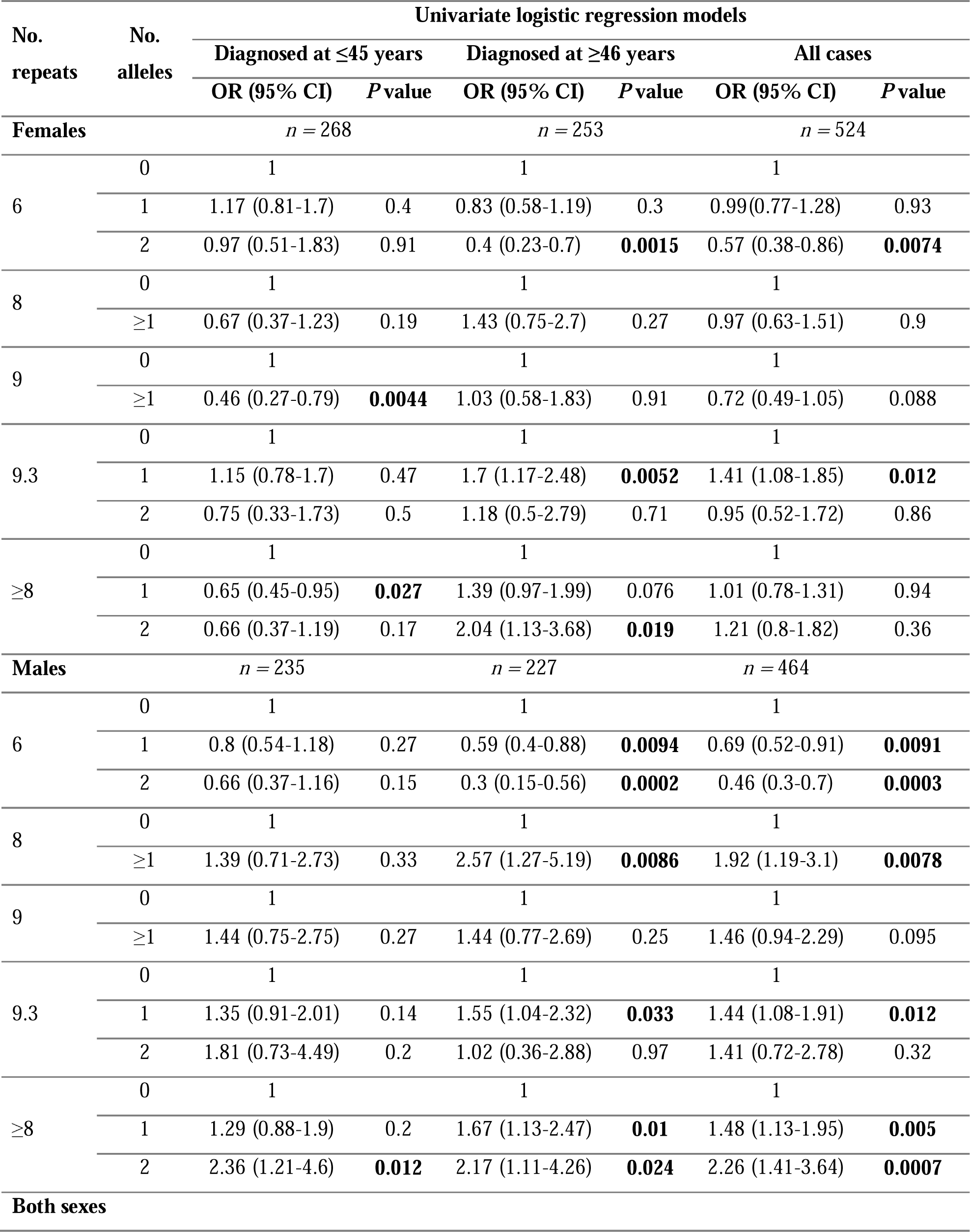
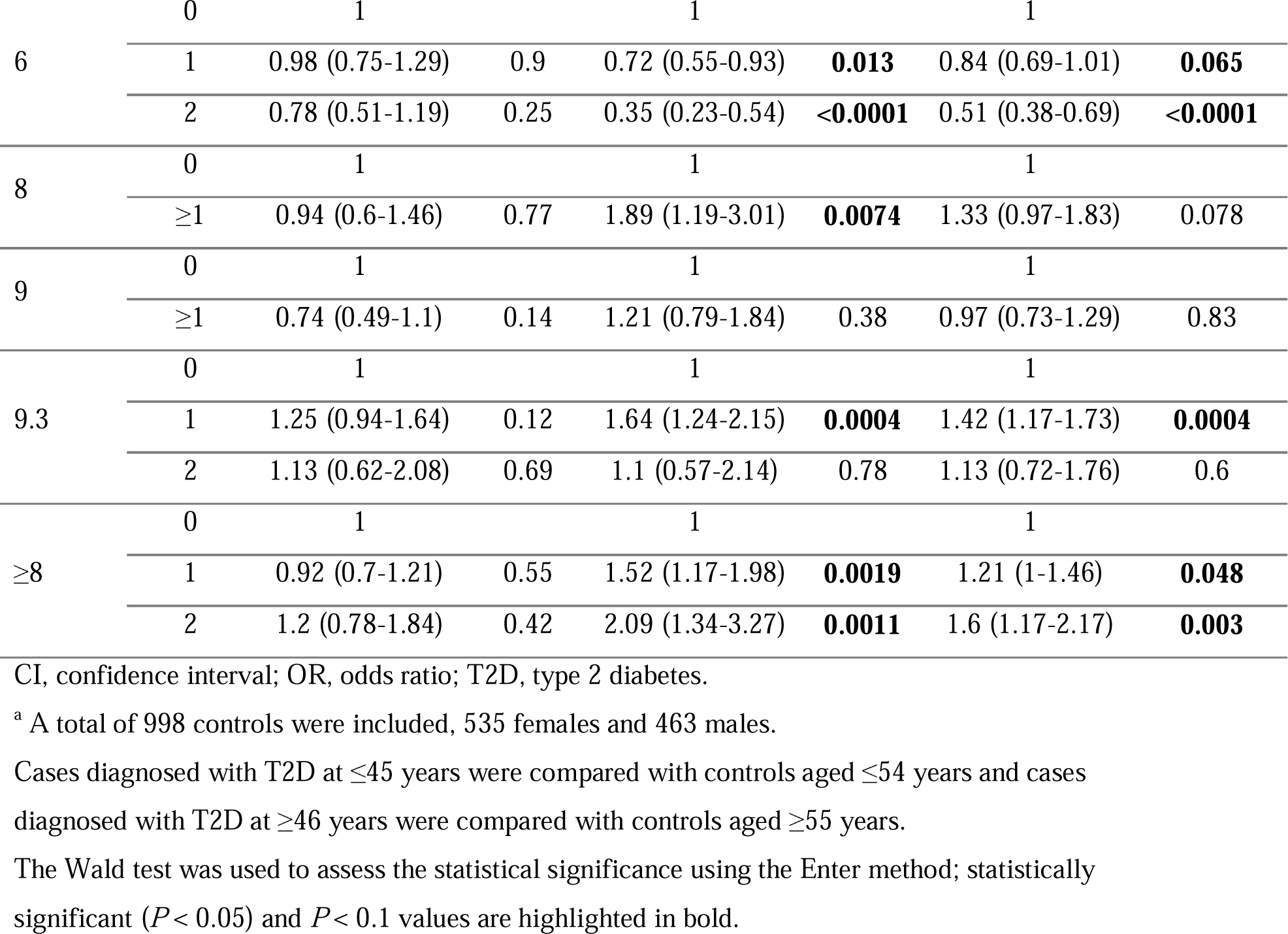
Association of *TH01* genotypes with T2D stratified by age at T2D diagnosis and sex (*n =* 1986)^a^.

### Frequency of alleles and genotypes of SNP rs689 and its association with T2D

**Table 3** compares the frequency of rs689 alleles (*n =* 8682) between the controls and cases. Allele A was significantly more frequent in cases than in controls (23.8% vs 20.0%; *P* < 0.0001). Although this difference was observed in females, it was markedly greater among males. This difference also significantly increased in male cases diagnosed at ≤45 years than at ≥46 years. In contrast, the frequency of allele A was also higher in cases than in controls among females diagnosed at ≥46 years, but not in the younger (≤45 years) group. The frequency of rs689 genotypes (*n =* 4341) between controls and cases are listed in **Table S3**, and similar differences were found in the frequency of genotypes AT and AA between all cases and controls.

**Table 3.** Comparison of frequency of rs689 alleles between controls and cases and stratified by age at type 2 diabetes diagnosis and sex (*n =* 8682).

| Allele | Allelic frequency, % ($n$) | | | | | |
| --- | --- | --- | --- | --- | --- | --- |
|  | Controls<br>≤54 years | Cases Dx<br>≤45 years | Controls<br>≥55 years | Cases Dx<br>≥46 years | All<br>controls | All cases |
| **Females** | $n = 2060$ | $n = 866$ | $n = 1434$ | $n = 914$ | $n = 3494$ | $n = 1780$ |
| **T** | 79.4 (1635) | 78.1 (676) | 80.3 (1152) | 76.7 (701) | 79.8 (2787) | 77.4 (1377) |
| **A** | 20.6 (425) | 21.9 (190) | 19.7 (282) | **23.3 (213)<sup>b</sup>** | 20.2 (707) | **22.6 (403)<sup>b</sup>** |
| **Males** | $n = 1038$ | $n = 824$ | $n = 788$ | $n = 758$ | $n = 1826$ | $n = 1582$ |
| **T** | 80.7 (838) | 73.7 (607) | 80.1 (631) | **76.4 (579)** | 80.4 (1469) | 75 (1186) |
| **A** | 19.3 (200) | **26.3 (217)<sup>d</sup>** | 19.9 (157) | **23.6 (179)<sup>a</sup>** | 19.6 (357) | **25 (396)<sup>d</sup>** |
| **Both sexes** | $n = 3098$ | $n = 1690$ | $n = 2222$ | $n = 1672$ | $n = 5320$ | $n = 3362$ |
| **T** | 79.8 (2473) | 75.9 (1283) | 80.2 (1783) | 76.6 (1280) | 80.0 (4256) | 76.2 (2563) |
| **A** | 20.2 (625) | **24.1 (407)<sup>c</sup>** | 19.8 (439) | **23.4 (392)<sup>c</sup>** | 20.0 (1064) | **23.8 (799)<sup>d</sup>** |

Dx, age at type 2 diabetes diagnosis.
<sup>a</sup> $P < 0.1$ ; <sup>b</sup> $P < 0.05$ ; <sup>c</sup> $P < 0.01$ ; <sup>d</sup> $P < 0.0001$ .
The Pearson chi square test was used to assess statistical significance; statistically significant ( $P < 0.05$ ) and $P < 0.1$ values are highlighted in bold.

Logistic regression analysis demonstrated that allele A confers a significantly higher risk for T2D (OR, 1.25; 95% CI, 1.1–1.4; *P* < 0.0001), with a comparatively higher risk in males (OR, 1.37; 95% CI, 1.2–1.6; *P* < 0.001) than in females (OR, 1.15; 95% CI, 1–1.3; *P* = 0.043). This risk was even higher in males diagnosed with T2D at ≤45 years (OR, 1.45; 95% CI, 1.2–1.9, *P* < 0.001) than at ≥46 years (OR, 1.25; 95% CI, 1–1.6; *P =* 0.069). This allele confers an increased risk for T2D only in females diagnosed at ≥46 years (OR, 1.24; 95% CI, 1–1.5; *P* = 0.032).

The association of SNP rs689 genotypes (*n* = 4341) are shown in **Table 4**. Both genotypes were found to confer a risk for T2D only in males diagnosed early, with an additive effect; risk was much greater for AA (OR, 2.2; 95% CI, 1.2–4.1; *P* = 0.013) than AT (OR, 1.53; 95% CI, 1.2–2; *P* = 0.002). In those diagnosed at ≥46 years, a higher risk was conferred only by AA in females (OR, 2.44; 95% CI, 1.4–4.2; *P =* 0.0015) and AT in males (OR, 1.38; 95% CI, 1–1.9; *P =* 0.036).

**Table 4.**
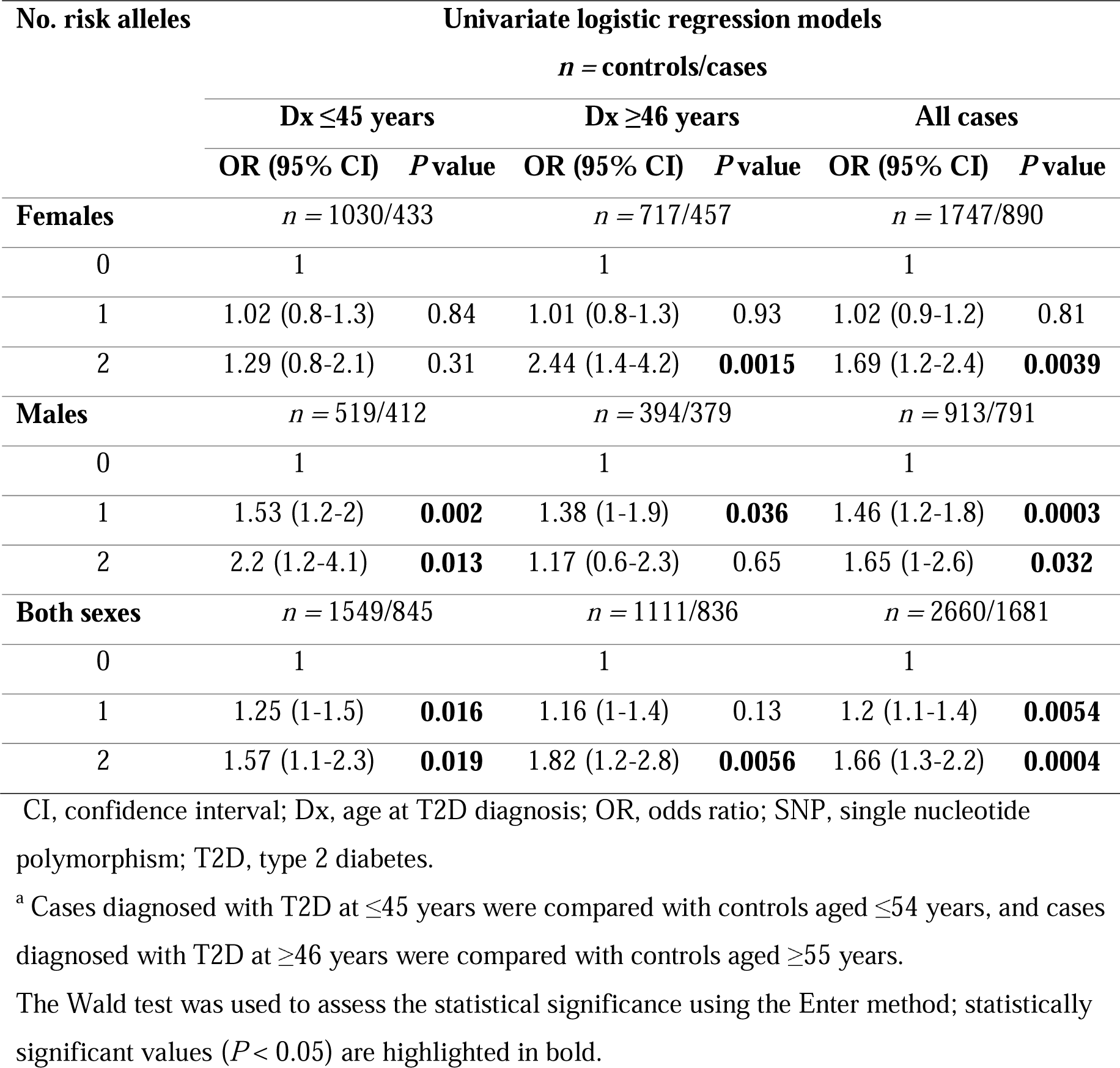
Association of SNP rs689 genotypes stratified by age at T2D diagnosis and sex (*n =* 4341)^a^.

The allelic and genotypic frequencies of SNP rs689 alleles (*n =* 3962) and genotypes (*n =* 1981) in the main study are presented in **Table S4**, and their associations using regression models in **Table S5**. Similar observations in the replica case–control study (alleles, *n =* 2376; genotypes *n =* 1188) are shown in **Table S6**, and their associations using regression models in **Table S7**; those for the cross-sectional study (alleles, *n =* 2344; genotypes, *n =* 1172) are in **Table S8** and **Table S9**, respectively. Despite the similar differences across the three studies in the allelic and genotypic frequencies, the risk conferred by allele A, or genotypes AT and AA, towards T2D diagnosis were observed mainly in males diagnosed at an earlier age in the two replica studies.

### Linkage disequilibrium analysis

The linkage disequilibrium between the *TH01* and SNP rs689 alleles (*n =* 3682) is summarized in **Table 5**. Interestingly, the alleles of *TH01* with 3 to 7 repeats were in linkage disequilibrium with allele T of SNP rs689. Alleles 6 and 7 of *TH01* were inherited with allele T of SNP rs689 98.5% (Tajima’s D [D’] = 0.9307) and 94.8% (D’ = 0.7585) of the time, respectively. The differences in the frequency of haplotypes 6/T vs 6/A and 7/T vs 7/A completely departed from the expected random distribution if they were in linkage equilibrium (*P <* 0.00001). In contrast, all *TH01* alleles with 8 to 11 repeats, except allele 9, were in partial linkage disequilibrium with allele A of SNP rs689. Allele 9.3 of *TH01* was highly linked and inherited with allele A for 83.0% of the time (D’ = 0.7829), whereas alleles 8 and 11 were partially linked to allele A and segregated together for 52.9% and 66.7% of the time, respectively, and their respective D’ values were lower (0.3882 and 0.5753). Similarly, the frequencies of haplotypes 9.3/A, 8/A, and 11/A were much more frequent than haplotypes with a T than if they were in equilibrium and randomly distributed (*P <* 0.01). Allele 9 was in linkage disequilibrium with allele T, and they were inherited together 90.1% of the time (D’ = 0.5814).

**Table 5.** Linkage disequilibrium between *TH01* and SNP rs689 alleles (*n =* 3682).

| *TH01*<br>alleles | Frequency of SNP rs689 alleles |  |  | Disequilibrium Linkage |  |  |  |
| --- | --- | --- | --- | --- | --- | --- | --- |
| | Observed/Expected | | Observed<br>T+A | *P* value | D' | | $r^2$ |
|  | T | A |  |  | T | A |  |
| **3** | 4/3 | 0/1 | 4 | >0.05 | 1 | −1 | 0.0003 |
| **4** | 2/2 | 0/0 | 2 | >0.05 | 1 | −1 | 0.0001 |
| **5** | 2/2 | 0/0 | 2 | >0.05 | 1 | −1 | 0.0001 |
| **6** | 1190/948 | 18/260 | 1208 | <0.0001 | **0.9307** | −0.9307 | 0.1159 |
| **7** | 1296/1073 | 71/294 | 1367 | <0.0001 | **0.7585** | −0.7585 | 0.0931 |
| **8** | 81/135 | 91/37 | 172 | <0.0001 | −0.4 | **0.4** | 0.0286 |
| **9** | 192/166 | 19/45 | 211 | <0.0001 | **0.5814** | −0.5814 | 0.0056 |
| **9.3** | 121/557 | 589/153 | 710 | <0.0001 | −0.7829 | **0.7829** | 0.5343 |
| **11** | 2/5 | 4/1 | 6 | 0.0071 | −0.5753 | **0.5753** | 0.0020 |
| **Total** | 2890 | 792 | 3682 |  |  |  |  |

D', Tajima's D; SNP, single nucleotide polymorphism.
Statistically significant values ( $P < 0.05$ ) are highlighted in bold.

### Multivariate logistic regression of TH01 and SNP rs689

The associations of the *TH01* and SNP rs689 alleles (*n =* 3952) with T2D are shown in **Table 6**. Both markers were associated with T2D in the same groups as reported in ULR models; the association remained significant for both loci in MLR models. Despite being in linkage disequilibrium, the ligation between some alleles of these two markers is incomplete, suggesting that both loci contribute independently to T2D in the MLR model. In fact, the value of R^2^ increased from the first (*TH01*) to the second (rs689) block introduced in the MLR model, supporting the additive contribution of the second marker in the model, which could be indicative of a cooperative effect by which both loci contribute to T2D.

**Table 6.**
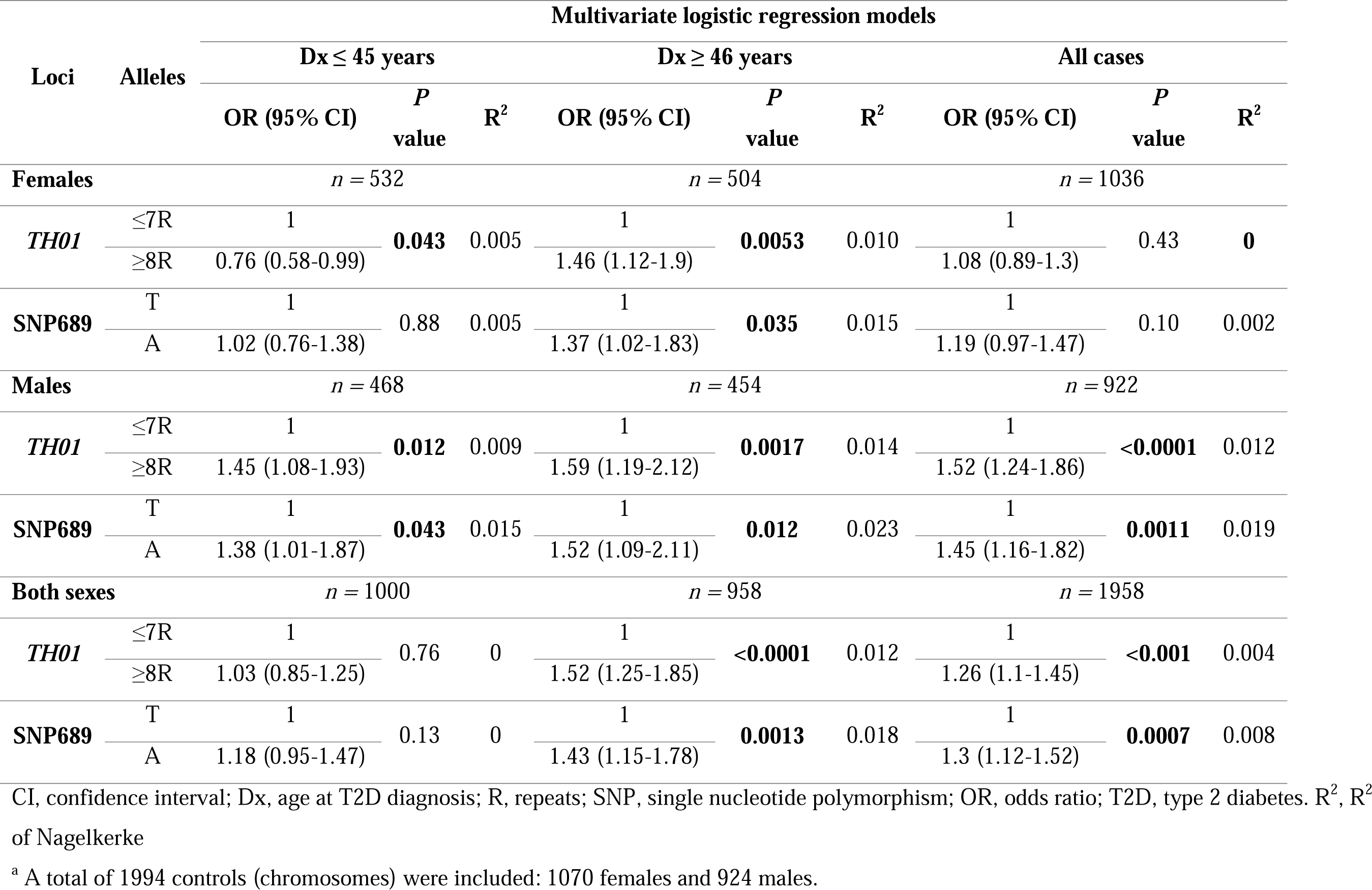
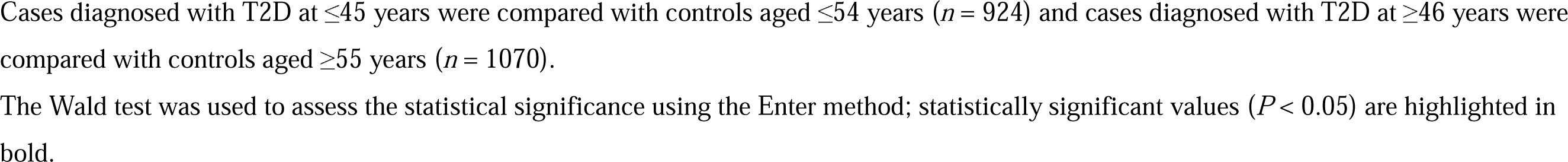
Association of *TH01* and SNP rs689 alleles with T2D stratified by age at T2D diagnosis and sex (*n =* 3952)^a^.

| Loci | Alleles | Multivariate logistic regression models |  |  |  |  |  |  |  |  |
| --- | --- | --- | --- | --- | --- | --- | --- | --- | --- | --- |
|  |  | Dx ≤ 45 years |  |  | Dx ≥ 46 years |  |  | All cases |  |  |
|  |  | OR (95% CI) | *P* value | R<sup>2</sup> | OR (95% CI) | *P* value | R<sup>2</sup> | OR (95% CI) | *P* value | R<sup>2</sup> |
| Females |  | *n* = 532 |  |  | *n* = 504 |  |  | *n* = 1036 |  |  |
| *TH01* | ≤7R | 1 | **0.043** | 0.005 | 1 | **0.0053** | 0.010 | 1 | 0.43 | **0** |
|  | ≥8R | 0.76 (0.58-0.99) |  |  | 1.46 (1.12-1.9) |  |  | 1.08 (0.89-1.3) |  |  |
| SNP689 | T | 1 | 0.88 | 0.005 | 1 | **0.035** | 0.015 | 1 | 0.10 | 0.002 |
|  | A | 1.02 (0.76-1.38) |  |  | 1.37 (1.02-1.83) |  |  | 1.19 (0.97-1.47) |  |  |
| Males |  | *n* = 468 |  |  | *n* = 454 |  |  | *n* = 922 |  |  |
| *TH01* | ≤7R | 1 | **0.012** | 0.009 | 1 | **0.0017** | 0.014 | 1 | <**0.0001** | 0.012 |
|  | ≥8R | 1.45 (1.08-1.93) |  |  | 1.59 (1.19-2.12) |  |  | 1.52 (1.24-1.86) |  |  |
| SNP689 | T | 1 | **0.043** | 0.015 | 1 | **0.012** | 0.023 | 1 | **0.0011** | 0.019 |
|  | A | 1.38 (1.01-1.87) |  |  | 1.52 (1.09-2.11) |  |  | 1.45 (1.16-1.82) |  |  |
| Both sexes |  | *n* = 1000 |  |  | *n* = 958 |  |  | *n* = 1958 |  |  |
| *TH01* | ≤7R | 1 | 0.76 | 0 | 1 | <**0.0001** | 0.012 | 1 | <**0.001** | 0.004 |
|  | ≥8R | 1.03 (0.85-1.25) |  |  | 1.52 (1.25-1.85) |  |  | 1.26 (1.1-1.45) |  |  |
| SNP689 | T | 1 | 0.13 | 0 | 1 | **0.0013** | 0.018 | 1 | **0.0007** | 0.008 |
|  | A | 1.18 (0.95-1.47) |  |  | 1.43 (1.15-1.78) |  |  | 1.3 (1.12-1.52) |  |  |

CI, confidence interval; Dx, age at T2D diagnosis; R, repeats; SNP, single nucleotide polymorphism; OR, odds ratio; T2D, type 2 diabetes. R<sup>2</sup>, R<sup>2</sup> of Nagelkerke
<sup>a</sup> A total of 1994 controls (chromosomes) were included: 1070 females and 924 males.
Cases diagnosed with T2D at $\leq 45$ years were compared with controls aged $\leq 54$ years ( $n = 924$ ) and cases diagnosed with T2D at $\geq 46$ years were compared with controls aged $\geq 55$ years ( $n = 1070$ ).
The Wald test was used to assess the statistical significance using the Enter method; statistically significant values ( $P < 0.05$ ) are highlighted in bold.
CI, confidence interval

The association of *TH01*/rs689 haplotypes (*n =* 3952) with T2D, using the univariate model, is shown in **Table S10**. In males, both haplotypes were associated with T2D with haplotype ≥8R/T conferring a higher risk than haplotype ≥8R/A. This finding indicates a predominant effect of the microsatellite *TH01* over that of rs689 alleles. When explored individually, the T allele of SNP rs689 confers protection, although in females the effect differed between the age groups. Here, in females ≥8R/T was protective for T2D development at ≤45 years while ≥8R/A had no effect; the opposite effect was observed among the group diagnosed with T2D at ≥46 years of age.

### Association of insulin concentration with age and the alleles of TH01 and rs689

**Table S11** depicts the concentrations of fasting plasma insulin observed in the main study, as stratified by age at T2D diagnosis, sex, and *TH01* and rs689 alleles (*n =* 2564). The median (interquartile range) fasting plasma insulin concentration (mIU/mL) was significantly higher among cases than in controls: 9.5 (5.5–16.2) vs 6.8 (4.8–10); *P <* 0.0001. Interestingly, insulin concentrations were higher in cases diagnosed at ≤45 years than at ≥46 years; this difference was greater in males than in females. In fact, insulin concentrations decreased with the age at T2D diagnosis (r = −0.111; *P <* 0.0001). The decrease was much greater in males (r = −0.189; *P <* 0.0001) than female (r = −0.051; *P* > 0.05) and was independent of allele type, suggesting it is likely related to age.

Insulin concentration significantly decreased with participant age for the entire main study population (r = −0.046; *P =* 0.02). The correlations between fasting plasma insulin concentration and age of controls and cases for the alleles of the two markers are shown for males in **Figure 2** and females in **Figure 3**. In males with diabetes, insulin decreased significantly with age in those with alleles that had ≥8R of *TH01* (r = −0.203; *P =* 0.009) or the rs689 allele A (r = −0.250; *P =* 0.004). With the presence of both alleles, there was a significant increase in the negative correlation (r = −0.423; *P =* 0.007), which occurred only with ≥8R/A and not ≥8R/T. On the contrary, in males in the control group, an inverse correlation was observed for those who have these alleles whereby the fasting plasma insulin concentration increased with age (**Figure 2**). In contrast, no significant correlations between insulin concentration and age were observed in cases with T2D or females in the control group for the alleles of the two markers (**Figure 3**).

**Figure 2.**
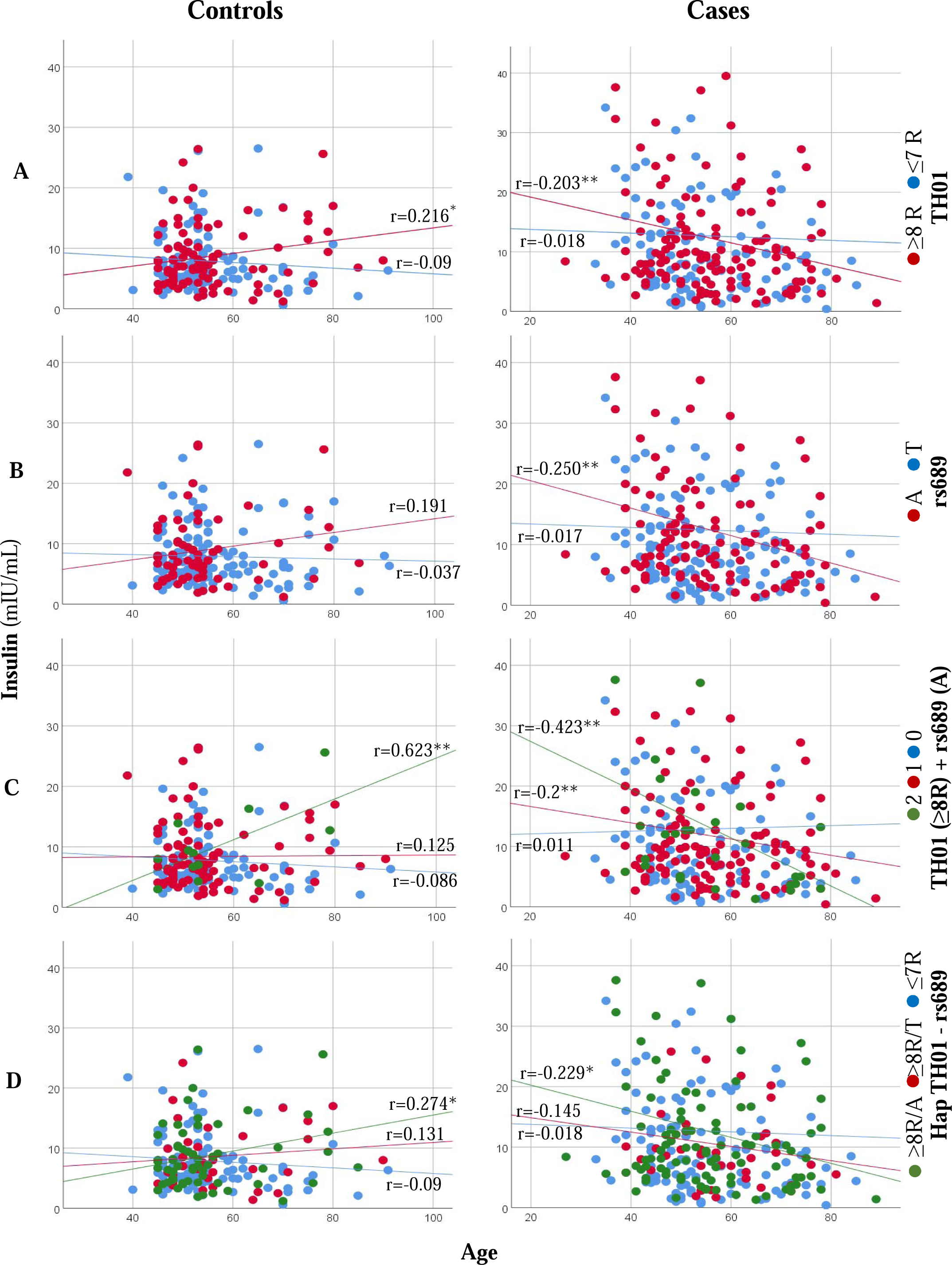
Correlation between fasting plasma insulin concentration and age of controls and cases in males for (A) *TH01* alleles (B) SNP rs689 alleles (C) *TH01* ≥8R alleles +/or rs689 A allele and (D) Hap *TH01* – rs689. In panel C, the numbers 2 (green circle), 1 (red circle), and 0 (blue circle) represent *TH01* (≥8R) + rs689 A allele, *TH01* (≥8R) or rs689 A allele, and neither, respectively. *p<0.05, **p<0.01. R, repeats; SNP, single nucleotide polymorphism.

**Figure 3.**
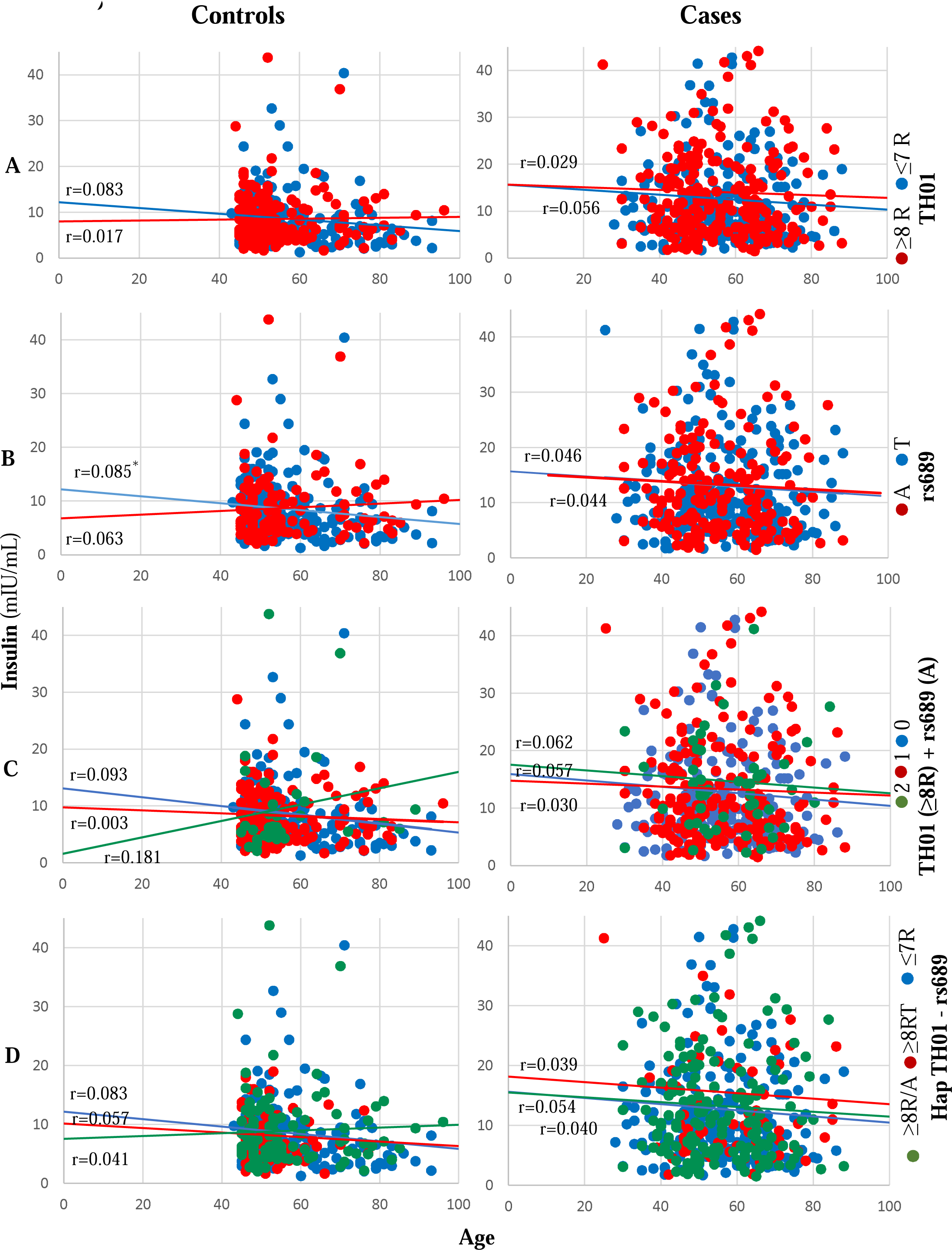
Correlation between fasting plasma insulin concentration and age of controls and cases in females for (A) *TH01* alleles (B) SNP rs689 alleles (C) *TH01* ≥8R alleles +/or rs689 A allele and (D) Hap *TH01* – rs689. In panel C, the numbers 2 (green circle), 1 (red circle), and 0 (blue circle) represent *TH01* (≥8R) + rs689 A allele, *TH01* (≥8R) or rs689 A allele, and neither, respectively. *p<0.05, **p<0.01. R, repeats; SNP, single nucleotide polymorphism.

## DISCUSSION

Our results indicate an association between *TH01* microsatellite and SNP rs689 with T2D and with plasma concentrations of fasting plasma insulin. The degree of association varied with age at T2D diagnosis and sex. *TH01* alleles with ≥8R and rs689 A allele conferred an increased risk of developing T2D at any age in males and at ≥46 years in females; there was a protective effect for developing T2D at ≤45 years among females. *TH01* alleles with ≤7R have either an inverse (allele 6) or a neutral (allele 7) effect. Fasting plasma insulin decreased linearly with age in male cases who had alleles with ≥8R or rs689 A allele but increased in controls with those alleles. In contrast, fasting plasma insulin remained constant in male cases and controls who had alleles with ≤7R or rs689 T allele. These findings are in line with those reported by Le Stunff et al who found that, when using SNP rs689 as a surrogate marker of *INS* VNTR, T/T genotypes (VNTR I/I) were associated with a higher level of fasting plasma insulin [36]. On the contrary, these results suggest that the decrease of insulin concentration with age in females is not influenced by *TH01* or rs689.

Linkage disequilibrium and multivariate analysis suggest that the *TH01* marker is partially in linkage imbalance with SNP rs689 and that the risk or protection conferred by both loci for T2D appear to be independent of each other, even though there may be an additive effect. On analyzing SNP rs689 in isolation, the T allele conferred protection for T2D, but when linked to *TH01* alleles with ≥8R, the haplotype (≥8R/T) conferred a risk of developing T2D in males at any age and in females aged ≥46 years. However, alleles with ≥8R conferred protection for T2D in females aged ≤45 years when linked to the T allele (≥8R/T), but not the A allele (≥8R/A).

Although inconsistent, most studies indicate that class I alleles stimulate the *INS* gene 1.5- to 3-times more than class III alleles [10, 14, 17, 18, 37–39]. A study that used the SNP rs689 as a surrogate marker of the *INS* VNTR noted an association between T/T genotypes (VNTR I/I) with a higher fasting plasma insulin level [36]. In contrast, some studies have found greater stimulation of allele III for insulin gene expression in *in vitro* experiments [15] or equal to allele I for plasma insulin secretion [16]. Allele A of SNP rs689, linked with allele III, influences alternative splicing of intron 1 of *INS* through differential recognition of its 3′ splice site, resulting in an increased production of mature transcripts and more proinsulin in culture supernatants than transcripts from allele T [16].

The influence of VNTR on gene activity or plasma insulin secretion in T2D remains unknown. However, studies have demonstrated the involvement of VNTR in the expression of *INS*, a mechanism that could be altered or affected in T2D in some populations [11, 16]. Our results with SNP rs689 support this hypothesis, as insulin concentration significantly decreased with age in male participants with T2D who were positive for allele A (class III), while it remained constant in those positive for allele T (class I).

We found that alleles with ≥8R were associated with a slight increase in fasting plasma insulin concentrations with age in controls, indicating a probable involvement of *TH01* in regulating *INS* expression. However, because this association was also observed with allele A of SNP rs689, the association between *TH01* microsatellite alleles and fasting plasma insulin levels could be due to a linkage effect with allele A. In cases with these alleles or with the ≥8R/A haplotype, fasting plasma insulin levels decreased with age, which could suggest that the regulation of *INS* expression could be progressively affected by age in the presence of class III (of *VNTR INS*) and/or ≥8R (of *TH01*) alleles. Because allele 6, which is completely linked to allele T of rs689, was found to be a protector for T2D and its frequency increased with age, it could have an inverse effect compared with the ≥8R/A haplotype on *INS* gene expression during aging.

Given alleles with ≥8R are protective for T2D development at ≤45 years and confer a greater risk at a later age (only in females), then this could indicate the crucial role of estrogen in this association. Epidemiological studies have demonstrated that estrogens are protective for T2D, whereas hormone therapy has been shown to reduce the incidence of diabetes by 35% in postmenopausal females with coronary heart disease [40]. This is in contrast to early menopause, during which there is a higher risk of T2D [41]. There is evidence that estrogen represses the expression of insulin mRNA in pancreatic β-cells through indirect genomic signaling [42] and can stimulate the degradation of misfolded proinsulin, thereby protecting the production of insulin and delaying the onset of diabetes [43]. Our analysis is limited by including only Mexican participants and therefore the study findings are not necessarily generalizable to other populations. Moreover, this study focused on the diagnosis of T2D; however, the extent of it and whether the investigated alleles pose a risk for treatment failure, remain unknown.

In conclusion, insulin increases with age in males when *TH01* alleles with ≥8R or allele A of rs689 are present, suggesting an involvement in a mechanism that maintains insulin synthesis in individuals without T2D. In contrast, this is somehow disabled in patients with T2D, thus warranting further research.

## Materials and Methods

### Sample selection and study design

The individuals included in the main case–control study were part of the Diabetes in Mexico Study (DMS), the details of which have been previously described as part of the SIGMA Type 2 Diabetes Consortium [23, 24]. Briefly, participants were recruited between November 2009 and August 2013 from two tertiary level hospitals in Mexico City, Mexico; T2D was diagnosed based on the American Diabetes Association [44] criteria. A total of 988 cases (unrelated individuals, aged >20 years, with a previous diagnosis of T2D or fasting blood glucose >125 mg/dL) and 998 controls (healthy individuals aged >50 years with fasting blood glucose <100 mg/dL) were selected from the DMS database; all participants were of Mexican mestizo ancestry. Clinical information such as weight, height, waist and hip circumference, and parental history of T2D was collected during the initial interview. For fasting glucose measurements and DNA extraction, 10 mL of intravenous blood was collected; all participants were assessed for *TH01* and rs689 markers.

For the replication of rs689, a case–control study including 593 cases of T2D and 595 controls (recruited between January 2014 and January 2015 from a tertiary care hospital in Mexico City, Mexico) was designed. Cases were individuals previously diagnosed with T2D according to the American Diabetes Association criteria, who agreed to participate, and were continuously recruited during their routine medical visits. Controls were individuals aged ≥50 years, who attended the same clinics for reasons other than T2D, had fasting blood glucose <100 mg/dL, and agreed to participate in the study. Clinical history and anthropometric and biochemical measurements were recorded for all study participants.

For another replica of rs689, a population-based cross-sectional study was conducted in 2005 patients (recruited between July and December 2017 from a hospital in Puebla, Mexico). Healthy individuals were invited to participate through flyers distributed in the hospital’s neighborhood. The participants were surveyed to assess T2D risk factors, and anthropometric measurements were collected. A blood sample was drawn to measure glycated hemoglobin (A1c) and assess DNA polymorphisms; individuals with A1c ≥6.5 were considered diabetic, and those with A1c <6.5 were considered non-diabetic. The SNP, rs689 was genotyped in all patients with T2D (*n* = 104), those with pre-diabetes (*n* = 529; A1c ≥5.7 to <6.5), and controls aged ≥40 years (*n* = 539; A1c <5.7). Those with pre-diabetes and controls were included in the non-diabetic group (*n* = 1068).

The associations of *TH01* microsatellite and rs689 were investigated using linear regression models. For the analysis, the presence or absence of T2D was considered the dependent variable, and the values of the alleles or genotypes were considered the explanatory variables.

### Ethics

The protocol for the main study was approved by the local ethics committee of each study site and the Federal Commission for the Protection Against Health Risks (COFEPRIS) (CAS/OR/CMN/113300410D0027-0577/2012). The clinical replica study was approved by the Ethics and Research Committees of the Comisión Nacional de Investigación Científica of the Instituto Mexicano del Seguro Social (IMSS R-2014-785-005), whilst the cross-sectional study was approved by the Ethics and Research Committees of the Hospital General de Puebla “Ignacio Romero Vargas” (68/ENS/INV/REV/2017). All three protocols complied with the Declaration of Helsinki and local ethical guidelines for clinical studies in Mexico. Written informed consent was obtained from all participants prior to their inclusion in the study.

### TH01 and rs689 SNP genotyping

All DNA samples of the main study were genotyped for *TH01* and all samples from the replica studies were genotyped for SNP rs689. The *TH01* microsatellite was assessed by polymerase chain reaction (PCR) using a fluorescence-labelled primer (5′- GTGGGCTGAAAAGCTCCCGATTAT-3′) and an unlabeled primer (5′- ATTCAAAGGGTATCTGGGCTCTGG-3′). Subsequently, alleles were identified by number of repeats (R) identified during capillary electrophoresis using the SeqStudio Genetic Analyzer (ThermoFisher Scientific, Waltham, MA, USA). Genotyping of the rs689 SNP was performed using the allelic discrimination assay-by-design TaqMan^®^ method (C_1223317_10) on 384-well plates analyzed on the QuantStudio™ 12 K Flex Real-Time PCR System (ThermoFisher Scientific). The genotypes were analyzed using Genotyper^™^ software v1.3 (ThermoFisher Scientific).

### Statistical analyses

For the sample size calculation, MLR models were considered to have good performance when a baseline of 100 cases (i.e., the ULR models) and 10–15 additional cases for each variable were introduced into the model [45]. Given that only two variables were to be introduced in the MLR models, we calculated that a minimum number of 115 cases were needed in the comparison groups of the main case–control study in which both loci were studied.

Baseline characteristics and insulin concentration were summarized using mean ± SD or median and interquartile range (25%–75%). To assess the statistical significance of intergroup differences, the Student’s *t-*test was used for mean values, and the Mann–Whitney *U* test for median values. We investigated whether the frequency of genotypes was distributed according to the Hardy–Weinberg law based on the allelic frequency and the formula: (*a* + *b*)^2^ = *a*^2^ + 2*ab* + *b*^2^, where *a* and *b* are the allelic frequencies in the control group. The statistical significance of the differences in the distribution of genotypes between the observed and expected results was calculated using the chi-square test, and the linkage disequilibrium between the *TH01* and rs689 SNP alleles was calculated using Arlequin (version 3.5.2.2; http://cmpg.unibe.ch/software/arlequin35/) [46]. The trend of fasting plasma insulin concentration was analyzed according to participant age; significance was calculated using the Pearson correlation test.

Univariate and multivariate logistic regression models were used to calculate the risk or protection conferred by the *TH01* microsatellite and VNTR loci for T2D development. The analysis of both DNA polymorphism variables was stratified by sex and age at T2D diagnosis. For these subgroups, cases (≤45 and ≥46 years) and controls (≤54 and ≥55 years) were categorized by their median age at diagnosis. The risk conferred by each factor (explanatory variables) was calculated by comparing the cases and controls using univariate logistic regression models. The reference indicators of the explanatory variables *TH01* and rs689, were *TH01* alleles with ≤7R and rs689 = T. The association was expressed as the OR and 95% CI, and the contribution to the variability of T2D was expressed as Nagelkerke’s R^2^. The factors were included successively in the model in different blocks. The contribution of each factor to the model was assessed by the increase of R^2^ and the decrease in the −2 log likelihood ratio value from one block to the next; the Omnibus test was used to determine statistical significance between the successive blocks.

A post-hoc power analysis was performed for each logistic regression model using the G*Power software version 3.1.9.7 (https://www.psychologie.hhu.de/arbeitsgruppen/allgemeine-psychologie-und-arbeitspsychologie/gpower), and when considering the sample size, OR, the probability of the event in the control group, and an α = 0.05 [47]. In addition, for multivariate linear regression models, the value of the total r^2^ obtained at the end of the model was introduced for the power calculation. All statistical tests were two-sided, and the level of significance was set at *P* < 0.05. The statistical analyses were conducted using SPSS software version 25 (IBM Corp., Armonk, NY, USA).

## Funding

This work was supported by the Carlos Slim Foundation, the Laboratorio Huella Génica, and the Faculty of Medicine of the National Autonomous University of Mexico (UNAM).

## Supporting information

STROBE Checklist

## Data Availability

All data produced in the present study are available upon reasonable request to the authors.

## Acknowledgements

The authors wish to thank Aafreen Saiyed of Edanz (www.edanz.com) for providing editorial support for this manuscript, in accordance with Good Publication Practice (GPP3) guidelines (http://www.ismpp.org/gpp3).

## Conflict of Interest Statement

All authors have nothing to disclose.

## Abbreviations

A1c,: glycated hemoglobin
CI: confidence interval
D’: Tajima’s D
DMS: Diabetes in Mexico Study
INS: insulin
L: large
MLR: multivariate logistic regression
OR: odds ratio
R: repeats
S: small
SD: standard deviation
SNP: single nucleotide polymorphism
T2D: type 2 diabetes
ULR: univariate logistic regression
VNTR: variable number of tandem repeats

## Notes

### Competing Interest Statement

The authors have declared no competing interest.

### Funding Statement

This work was funded by the Carlos Slim Foundation, the Laboratorio Huella Genica, and the Faculty of Medicine of the National Autonomous University of Mexico (UNAM).

### Author Declarations

The protocol for the main study was approved by the local ethics committee of each study site and the Federal Commission for the Protection Against Health Risks (COFEPRIS) (CAS/OR/CMN/113300410D0027-0577/2012). The clinical replica study was approved by the Ethics and Research Committees of the Comisión Nacional de Investigación Científica of the Instituto Mexicano del Seguro Social (IMSS R-2014-785-005), whilst the cross-sectional study was approved by the Ethics and Research Committees of the Hospital General de Puebla “Ignacio Romero Vargas” (68/ENS/INV/REV/2017). All three protocols complied with the Declaration of Helsinki and local ethical guidelines for clinical studies in Mexico.

## References

1. Kahn, S.E. (2001) The importance of beta-cell failure in the development and progression of type 2 diabetes. J. Clin. Endocrinol. Metab., 86, 4047–4058.

2. Quirós, R.Y., Lardoeyt, F.R., Arrieta, G.R., Medina, A.F.E. (2011) Influence of the genome-environment interaction in type 2 diabetes mellitus in a municipality of Camagüey province. Rev. Cub. Gen., 5, 13. Spanish.

3. Andersson, S.A., Olsson, A.H., Esguerra, J.L., Heimann, E., Ladenvall, C., Edlund, A., Salehi, A., Taneera, J., Degerman, E., Groop, L., et. al. (2012) Reduced insulin secretion correlates with decreased expression of exocytotic genes in pancreatic islets from patients with type 2 diabetes. Mol. Cell. Endocrinol., 364, 36–45.

4. Kralovicova, J. and Vorechovsky, I. (2010) Allele-specific recognition of the 3’ splice site of INS intron 1. Hum. Genet., 128, 383–400.

5. Polonsky, K.S., Given, B.D., Hirsch, L.J., Tillil, H., Shapiro, E.T., Beebe, C., Frank, B.H., Galloway, J.A. and Van Cauter, E. (1988) Abnormal patterns of insulin secretion in non-insulin-dependent diabetes mellitus. N. Engl. J. Med., 318, 1231–1239.

6. Noormets, K., Koks, S., Muldmaa, M., Mauring, L., Vasar, E. and Tillmann, V. (2011) Sex differences in the development of diabetes in mice with deleted wolframin (Wfs1) gene. Exp. Clin. Endocrinol. Diabetes, 119, 271–275.

7. Groenewoud, M.J., Dekker, J.M., Fritsche, A., Reiling, E., Nijpels, G., Heine, R.J., Maassen, J.A., Machicao, F., Schäfer, S.A., Häring, H.U. and t Hart, L.M. (2008) Variants of CDKAL1 and IGF2BP2 affect first-phase insulin secretion during hyperglycaemic clamps. Diabetologia, 51, 1659–1663.

8. Zhou, Y., Park, S.Y., Su, J., Bailey, K., Ottosson-Laakso, E., Shcherbina, L., Oskolkov, N., Zhang, E., Thevenin, T., Fadista, J. and Bennet, H. (2014) TCF7L2 is a master regulator of insulin production and processing. Hum. Mol. Genet., 23, 6419–31.

9. Berumen, J., Orozco, L., Betancourt-Cravioto, M., Gallardo, H., Zulueta, M., Mendizabal, L., Simon, L., Benuto, R.E., Ramírez-Campos, E., Marin, M., et. al. (2019) Influence of obesity, parental history of diabetes, and genes in type 2 diabetes: A case-control study. Sci. Rep., 9, 2748.

10. Bennett, S.T., Lucassen, A.M., Gough, S.C.L., Powell, E.E., Undlien, D.E., Pritchard, L.E., Merriman, M.E., Kawaguchi, Y., Dronsfield, M.J., Pociot, F. et. al., (1995) Susceptibility to human type 1 diabetes at IDDM2 is determined by tandem repeat variation at the insulin gene minisatellite locus. Nat. Genet., 9, 284–292.

11. Pugliese, A., Zeller, M., Fernandez, A., Zalcberg, L.J., Bartlett, R.J., Ricordi, C., Pietropaolo, M., Eisenbarth, G.S., Bennett, S.T. and Patel, D.D. (1997) The insulin gene is transcribed in the human thymus and transcription levels correlated with allelic variation at the INS VNTR-IDDM2 susceptibility locus for type 1 diabetes. Nat. Genet., 15, 293–297.

12. Stead, J.D., Buard, J., Todd, J.A. and Jeffreys, A.J. (2000) Influence of allele lineage on the role of the insulin minisatellite in susceptibility to type 1 diabetes. Hum. Mol. Genet., 9, 2929–2935.

13. Coleta, R.D., Alexander, A.L., Alva, C.B.D., Pinto, E.M., Billerbeck, A.E.C., Pachi, P.R., Longui, C.A., Garcia, R.M., Boguszewski, M., Ivo, J.P., Arnhold, I.J.O., Mendonca, B.B. and Costa, E.M.F. (2013) Insulin-like growth factor 1 gene (CA)n repeats and a variable number of tandem repeats of the insulin gene in Brazilian children born small for gestational age. Clinics (Sao Paulo), 68, 785–791.

14. Cocozza, S., Riccardi, G., Monticelli, A., Capaldo, B., Genovese, S., Krogh, V., Celentano, E., Farinaro, E., Varrone, S. and Avvedimento, V.E. (1988) Polymorphism at the 5’ end flanking region of the insulin gene is associated with reduced insulin secretion in healthy individuals. Eur. J. Clin. Invest., 18, 582–586.

15. Kennedy, G.C., German, M.S. and Rutter, W.J. (1995) The minisatellite in the diabetes susceptibility locus IDDM2 regulates insulin transcription. Nat. Genet., 9, 293–298.

16. Ahmed, S., Bennett, S.T., Huxtable, S.J., Todd, J.A., Matthews, D.R. and Gough, S.C.L. (1999) INS VNTR allelic variation and dynamic insulin secretion in healthy adult non-diabetic Caucasian subjects. Diabet. Med., 16, 910–917.

17. Lucassen, A.M., Screaton, G.R., Julier, C., Elliott, T.J., Lathrop, M. and Bell, J.I. (1995) Regulation of insulin gene expression by the IDDM associated, insulin locus haplotype. Hum. Mol. Genet., 4, 501–506.

18. Vafiadis, P., Bennett, S.T., Colle, E., Grabs, R., Goodyer, C.G. and Polychronakos, C. (1996) Imprinted and genotype-specific expression of genes at the IDDM2 locus in pancreas and leucocytes. J. Autoimmun., 9, 397–403.

19. Knowler, W.C., Pettitt, D.J., Vasquez, B., Rotwein, P.S., Andreone, T.L. and Permutt, M.A. (1984) Polymorphism in the 5’ flanking region of the human insulin gene. Relationships with noninsulin-dependent diabetes mellitus, glucose and insulin concentrations, and diabetes treatment in the Pima Indians. J. Clin. Invest., 74, 2129–2135.

20. Huxtable, S.J., Saker, P.J., Haddad, L., Walker, M., Frayling, T.M., Levy, J.C., Hitman, G.A., O’Rahilly, S., Hattersley, A.T. and McCarthy, M.I. (2000) Analysis of parent-offspring trios provides evidence for linkage and association between the insulin gene and type 2 diabetes mediated exclusively through paternally transmitted class III variable number tandem repeat alleles. Diabetes, 49, 126–130.

21. Lindsay, R.S., Walker, J.D., Halsall, I., Hales, C.N., Calder, A.A., Hamilton, B.A., Johnstone, F.D. and Scottish Multicentre Study of Diabetes in Pregnancy, (2003) Insulin and insulin propeptides at birth in offspring of diabetic mothers. J. Clin. Endocrinol. Metab., 88, 1664–1671.

22. Hansen, S.K., Gjesing, A.P., Rasmussen, S.K., Glümer, C., Urhammer, S.A., Andersen, G., Rose, C.S., Drivsholm, T., Torekov, S.K., Jensen, D.P., et al., (2004) Large-scale studies of the HphI insulin gene variable-number-of-tandem-repeats polymorphism in relation to Type 2 diabetes mellitus and insulin release. Diabetologia, 47, 1079–87.

23. Estrada, K., Aukrust, I., Bjørkhaug, L., Burtt, N.P., Mercader, J.M., Garcia-Ortiz, H., Huerta-Chagoya, A., Moreno-Macias, H., Walford, G., Flannick, J. et al., (2014) Association of a low-frequency variant in HNF1A with type 2 diabetes in a Latino population. JAMA, 311, 2305–2314.

24. Williams, A.L., Jacobs, S.B., Moreno-Macías, H., Huerta-Chagoya, A., Churchhouse, C., Márquez-Luna, C., García-Ortíz, H., Gómez-Vázquez, M.J., Burtt, N.P., Aguilar-Salinas, C.A. et. al., (2014) Sequence variants in SLC16A11 are a common risk factor for type 2 diabetes in Mexico. Nature, 506, 97–101.

25. McGinnis, R.E. and Spielman, R.S. (1995) Insulin expression: is VNTR allele 698 really anomalous? Nat. Genet., 10, 378–380.

26. Awata, T. (1997) IDDM and variable number of tandem repeats (VNTR) in the 5’region of the insulin gene: a review. Nihon Rinsho, 55 Suppl, 376-381. Japanese.

27. Gymrek, M. (2017) A genomic view of short tandem repeats. Curr. Opin. Genet. Dev., 44, 9–16.

28. Rodriguez, S., Gaunt, T.R. and Day, I.N. (2007) Molecular genetics of human growth hormone, insulin-like growth factors and their pathways in common disease. Hum Genet., 122, 1–27.

29. Gu, D., O’Dell, S.D., Chen, X.H., Miller, G.J. and Day, I.N. (2002) Evidence of multiple causal sites affecting weight in the IGF2-INS-TH region of human chromosome 11. Hum. Genet., 110, 173–181.

30. Sharma, P., Hingorani, A., Jia, H., Ashby, M., Hopper, R., Clayton, D. and Brown, M.J. (1998) Positive association of tyrosine hydroxylase microsatellite marker to essential hypertension. Hypertension, 32, 676–682.

31. Jindra, A. (2000) Association analysis of two tyrosine hydroxylase gene polymorphisms in normotensive offspring from hypertensive families. Blood Press., 9, 250–254.

32. Klintschar, M., Immel, U.D., Stiller, D., Kleiber M. (2005) TH01, a tetrameric short tandem repeat locus in the tyrosine hydroxylase gene: Association with myocardial hypertrophy and death from myocardial infarction? Dis. Markers, 21, 9–13.

33. Rodriguez, S., Gaunt, T., ÓDell, S., Chen, X., Gu, D., Hawe, E., Miller, G., Humphries, S., Day, N. (2004) Haplotypic analyses of the IGF2-INS-TH gene cluster in relation to cardiovascular risk traits. Human Mol. Genet., 13, 715–725.

34. Rodriguez, S., Gaunt, T., Dennison, E., Chen, X., Syddall, E., Phyllips, D., Copper, S., Day, N. (2006) Replication of IGF2-INS-TH*5 haplotype effect on obesity in older men and study of related phenotypes. Eur. J. Human Gen., 14, 109–116.

35. Puers C., Hammond H.A., Jin L., Caskey T. and Schumm J.W. (1993). Identification of repeat sequence heterogeneity at the polymorphic short tandem repeat locus HUMTH01 (AATG) n and reassignment of alleles in population analysis by using a locus-specific allelic ladder. Am. J. Hum. Genet., 53: 953–958.

36. Le Stunff, C., Fallin, D., Schork, N.J. and Bougnères, P. (2000) The insulin gene VNTR is associated with fasting insulin levels and development of juvenile obesity. Nat. Genet., 26, 444–446.

37. Owerbach, D. and Gabbay, K.H. (1996) The search for IDDM susceptibility genes: the next generation. Diabetes, 45, 544–551.

38. Bennett, S.T., Wilson, A.J., Esposito, L., Bouzekri, N., Undlien, D.E., Cucca, F., Nisticò, L., Buzzetti, R., Bosi, E., Pociot, F., et. al. (1997) Insulin VNTR allele-specific effect in type 1 diabetes depends on identity of untransmitted paternal allele. Nat. Genet., 17, 350–352.

39. Luthman, H., Söderling[Barros, J., Persson, B., Engberg, C., Stern, I., Lake, M., Franzén, S.Å., Israelsson, M., Rådén, B., Lindgren, B., et. al. (1989) Human insulin-like growth-factor-binding protein. Low-molecular-mass form: protein sequence and cDNA cloning. Eur. J. Biochem., 180, 259–265.

40. Kanaya, A.M., Herrington, D., Vittinghoff, E., Lin, F., Grady, D., Bittner, V., Cauley, J.A. and Barrett-Connor, E. (2003) Glycemic effects of postmenopausal hormone therapy: the Heart and Estrogen/progestin Replacement Study. A randomized, double-blind, placebo-controlled trial. Ann. Intern. Med., 138, 1–9.

41. Brand, J.S., Van Der Schouw, Y.T., Onland-Moret, N.C., Sharp, S.J., Ong, K.K., Khaw, K.T., Ardanaz, E., Amiano, P., Boeing, H., Chirlaque, M.D., et. al. (2013) Age at menopause, reproductive life span, and type 2 diabetes risk: results from the EPIC-InterAct study. Diabetes Care, 36, 1012–1019.

42. Sekido, T., Nishio, S.I., Ohkubo, Y., Sekido, K., Kitahara, J., Miyamoto, T. and Komatsu, M. (2019) Repression of insulin gene transcription by indirect genomic signaling via the estrogen receptor in pancreatic beta cells. In Vitro Cell. Dev. Biol. Anim., 55, 226–236.

43. Xu, B., Allard, C., Alvarez-Mercado, A.I., Fuselier, T., Kim, J.H., Coons, L.A., Hewitt, S.C., Urano, F., Korach, K.S., Levin, E.R., et. al. (2018) Estrogens promote misfolded proinsulin degradation to protect insulin production and delay diabetes. Cell Rep., 24, 181–196.

44. American Diabetes Association. (2022) Standards of medical care in diabetes-2022 Abridged for primary care providers. Clin. Diabetes, 40,10–38.

45. Babyak, M.A. (2004) What you see may not be what you get: a brief, nontechnical introduction to overfitting in regression-type models. Psychosom. Med., 66, 411–421.

46. Excoffier, L. and Lischer, H.E.L. (2010) Arlequin suite ver 3.5: A new series of programs to perform population genetics analyses under Linux and Windows. Mol. Ecol. Resour., 10, 564–567.

47. Faul, F., Erdfelder, E., Lang, A.G. and Buchner, A. (2007) G*Power 3: a flexible statistical power analysis program for the social, behavioral, and biomedical sciences. Behav. Res. Methods, 39, 175–191.

