## Supplementary Tables 1-11 for "The *TH01* microsatellite and *INS* VNTR are strongly associated with type 2 diabetes and fasting insulin secretion"

**Supplementary Material**

**Table S1.** Participant demographic characteristics (*n* = 4346).

| Variable | Women | | Men | | Both sexes | |
| --- | --- | --- | --- | --- | --- | --- |
|  | **Control** | **Cases** | **Control** | **Cases** | **Control** | **Cases** |
| Main case–control study (*n =* 1986): Mean ± SD (*n*) | | | | | | |
| Age (years) | 59.7 ± 11.2 (535) | 55.4 ± 12.1 (524) | 58.6 ± 11.4 (463) | 55.8 ± 11.4 (464) | 59.2 ± 11.3 (998) | 55.6 ± 11.7 (988) |
| Age at diabetes diagnosis (years) |  | 45.9 ± 10.6 (524) |  | 46.2 ± 10.9 (464) |  | 46 ± 10.8 (988) |
| Years with the disease |  | 9.4 ± 8.4 (524) |  | 9.6 ± 9 (464) |  | 9.5 ± 8.7 (988) |
| Replica case–control study (*n =* 1188): Mean ± SD (*n*) | | | | | | |
| Age (years) | 53.4 ± 9.6 (423) | 57 ± 8.8 (297) | 55.8 ± 11 (172) | 57.3 ± 10.2 (296) | 54.1 ± 10.1 (595) | 57.1 ± 9.5 (593) |
| Age at diabetes diagnosis (years) |  | 45.3 ± 7 (297) |  | 44.3 ± 7.5 (296) |  | 44.8 ± 7.3 (593) |
| Years with the disease |  | 11.7 ± 7.8 (297) |  | 12.9 ± 7.5 (296) |  | 12.3 ± 7.7 (593) |
| Cross-sectional study (*n =* 1172): Mean ± SD (*n*) | | | | | | |
| Age (years) | 49.3 ± 10.7 (789) | 49 ± 11.7 (72) | 48.8 ± 11.1 (279) | 49.5 ± 10.9 (32) | 49.1 ± 10.8 (1068) | 49.2 ± 11.4 (104) |
| Age at diabetes diagnosis (years)^a^ |  | 49.8 ± 11.7 (63) |  | 48.8 ± 10.6 (29) |  | 49.5 ± 11.3 (92) |
| Years with the disease^b^ |  | ≈0 |  | ≈0.7 |  | ≈0 |

SD, standard deviation.

**^a^** Newly diagnosed cases were included and age at recruitment was used. The age at diabetes diagnosis of the 12 individuals with T2D diagnosed before recruitment was not collected.

**^b^** Difference between mean age of all cases less than the mean age of newly diagnosed cases.

**Table S2.** Association of *TH01* alleles with type 2 diabetes stratified by age at type 2 diabetes diagnosis and sex (*n* = 3972)^a^.

| Alleles^b^ | Univariate logistic regression models | | | | | |
| --- | --- | --- | --- | --- | --- | --- |
|  | **Controls ≤54 years vs. Cases dx ≤45 years** | | **Controls ≥55 years vs. Cases dx ≥46 years** | | **All Controls vs. All Cases** | |
|  | **OR (95% CI)** | ***P* value** | **OR (95% CI)** | ***P* value** | **OR (95% CI)** | ***P* value** |
| Women | *n =* 462/536 | | *n =* 608/506 | | *n =* 1070/1048 | |
| 0 | 1 |  | 1 |  | 1 |  |
| 6 | 1.05 (0.81-1.38) | 0.71 | **0.68 (0.53-0.87)** | **0.0023** | **0.83 (0.69-0.99)** | **0.039** |
| 0 | 1 |  | 1 |  | 1 |  |
| 8 | 0.73 (0.42-1.27) | 0.26 | 1.53 (0.84-2.78) | 0.17 | 1.04 (0.69-1.57) | 0.83 |
| 0 | 1 |  | 1 |  | 1 |  |
| 9 | **0.46 (0.28-0.77)** | **0.003** | 1.08 (0.62-1.87) | 0.79 | 0.72 (0.5-1.04) | 0.084 |
| 0 | 1 |  | 1 |  | 1 |  |
| 9.3 | 1.01 (0.74-1.38) | 0.94 | **1.43 (1.05-1.94)** | **0.021** | 1.21 (0.97-1.5) | 0.085 |
| ≤7R | 1 |  | **1** |  | 1 |  |
| ≥8R | **0.75 (0.58-0.98)** | **0.035** | **1.46 (1.12-1.91)** | **0.0049** | 1.07 (0.89-1.29) | 0.46 |
| Men | *n =* 464/470 | | *n =* 462/454 | | *n =* 926/928 | |
| 0 | 1 |  | 1 |  | 1 |  |
| 6 | 0.79 (0.6-1.04) | 0.096 | **0.56 (0.43-0.75)** | **<0.0001** | **0.67 (0.55-0.81)** | **<0.0001** |
| 0 | 1 |  | 1 |  | 1 |  |
| 8 | 1.44 (0.75-2.76) | 0.27 | **2.26 (1.18-4.33)** | **0.013** | **1.85 (1.17-2.91)** | **0.0084** |
| 0 | 1 |  | 1 |  | 1 |  |
| 9 | 1.41 (0.75-2.67) | 0.28 | 1.38 (0.77-2.47) | 0.28 | 1.42 (0.92-2.17) | 0.11 |
| 0 | 1 |  | 1 |  | 1 |  |
| 9.3 | 1.36 (0.98-1.88) | 0.062 | 1.33 (0.95-1.85) | 0.098 | **1.34 (1.06-1.69)** | **0.014** |
| ≤7R | 1 |  | 1 |  | **1** |  |
| ≥8R | **1.46 (1.1-1.95)** | **0.0088** | **1.57 (1.18-2.1)** | **0.0021** | **1.52 (1.24-1.86)** | **<0.0001** |
| Both sexes | *n =* 926/1006 | | *n =* 1070/960 | | *n =* 1996/1976 | |
| 0 | 1 |  | 1 |  | 1 |  |
| 6 | 0.91 (0.76-1.11) | 0.35 | **0.62 (0.52-0.75)** | **<0.0001** | **0.75 (0.66-0.86)** | **<0.0001** |
| 0 | 1 |  | 1 |  | 1 |  |
| 8 | 0.98 (0.64-1.5) | 0.93 | **1.85 (1.2-2.87)** | **0.0057** | **1.35 (1-1.83)** | **0.049** |
| 0 | 1 |  | 1 |  | 1 |  |
| 9 | 0.73 (0.49-1.07) | 0.11 | 1.22 (0.82-1.81) | 0.33 | 0.96 (0.73-1.27) | 0.78 |
| 0 | 1 |  | 1 |  | 1 |  |
| 9.3 | 1.17 (0.93-1.46) | 0.17 | **1.38 (1.1-1.73)** | **0.0048** | **1.27 (1.08-1.49)** | **0.0034** |
| ≤7R | 1 |  | 1 |  | **1** |  |
| ≥8R | 1.03 (0.85-1.25) | 0.75 | **1.52 (1.25-1.84)** | **<0.0001** | **1.26 (1.1-1.44)** | **0.0009** |

CI, confidence interval; OR, odds ratio; R, repeats.

Cases diagnosed with type 2 diabetes at ≤45 years were compared with controls aged ≤54 years and cases diagnosed with type 2 diabetes at ≥46 years were compared with controls aged ≥55 years. Statistically significant values are highlighted in bold (enter method, Wald test).

^a^ A total of 1996 controls (chromosomes) were included (female, *n* = 1070; male, *n* = 926).

^b^ 0 indicates the rest of alleles except the allele of the comparison.

**Table S3.** Comparison of frequency of rs689 genotypes between controls and cases of type 2 diabetes stratified by age at type 2 diabetes diagnosis and sex (*n =* 4341).

| Genotypes | Genotypic frequency % (*n*) | | | | | |
| --- | --- | --- | --- | --- | --- | --- |
|  | **Controls ≤54 years** | **Cases dx ≤45 years** | **Controls ≥55 years** | **Cases dx ≥46 years** | **All controls** | **All cases** |
| Women | *n =* 1030 | *n =* 433 | *n =* 717 | *n =* 457 | *n =* 1747 | *n =* 890 |
| TT | 63.5 (654) | 62.1 (269) | 63.9 (458) | **60.8 (278)** | 63.7 (1112) | **61.5 (547)** |
| AT | 31.7 (327) | 31.9 (138) | 32.9 (236) | **31.7 (145)** | 32.2 (563) | **31.8 (283)** |
| AA | 4.8 (49) | 6.0 (26) | 3.2 (23) | **7.4 (34)^c^** | 4.1 (72) | **6.7 (60)^a^** |
| AA/AT | 36.5 (376) | 37.9 (164) | 36.1 (259) | 39.2 (179) | 36.3 (635) | 38.5 (343) |
| Men | *n =* 519 | *n =* 412 | *n =* 394 | *n =* 379 | *n =* 913 | *n =* 791 |
| TT | 64.9 (337) | **53.6 (221)** | 64.7 (255) | 57.5 (218) | 64.8 (592) | **55.5 (439)** |
| AT | 31.6 (164) | **40.0 (165)** | 30.7 (121) | 37.7 (143) | 31.2 (285) | **38.9 (308)** |
| AA | 3.5 (18) | **6.3 (26)^c^** | 4.6 (18) | 4.7 (18) | 3.9 (36) | **5.6 (44)^c^** |
| AA/AT | 35.1 (182) | 46.4 (191) | 35.3 (139) | 42.5 (161) | 35.2 (321) | 44.5 (352) |
| Both sexes | *n =* 1549 | *n =* 845 | *n =* 1111 | *n =* 836 | *n =* 2660 | *n =* 1681 |
| TT | 64.0 (991) | **58.0 (490)** | 64.2 (713) | **59.3 (496)** | 64.1 (1704) | **58.7 (986)** |
| AT | 31.7 (491) | **35.9 (303)** | 32.1 (357) | **34.4 (288)** | 31.9 (848) | **35.2 (591)** |
| AA | 4.3 (67) | **6.2 (52)^c^** | 3.7 (41) | **6.2 (52)^b^** | 4.1 (108) | **6.2 (104)^c^** |
| AA/AT | 36.0 (558) | 42.0 (355) | 35.8 (398) | 40.7 (340) | 35.9 (956) | 41.3 (695) |

Statistically significant values are highlighted in bold (Pearson chi square test).

^a^ *P* <0.1; ^b^ *P* <0.05, ^c^ *P* <0.01, ^d^ *P* <0.001.

**Table S4.** Frequency of rs689 alleles (*n* = 3962) and genotypes (*n* = 1981) in the main case–control study stratified by sex and age of type 2 diabetes diagnosis.

|  | Controls ≤54 years | Cases Dx ≤45 years | Controls ≥55 years | Cases Dx ≥46 years | All controls | All cases |
| --- | --- | --- | --- | --- | --- | --- |
| Alleles | **Allelic frequency % (*n*)** | | | | | |
| Women | *n =* 462 | *n =* 532 | *n =* 608 | *n =* 504 | *n =* 1070 | *n =* 1042 |
| T | 78.1 (361) | 77.8 (414) | 81.7 (497) | 76.4 (385) | 80.2 (858) | 77.3 (805) |
| A | 21.9 (101) | 22.2 (118) | 18.3 (111) | **23.6 (119)^b^** | 19.8 (212) | 22.7 (237) |
| Men | *n =* 462 | *n =* 468 | *n =* 462 | *n =* 454 | *n =* 924 | *n =* 926 |
| T | 80.1 (370) | 74.4 (348) | 82.9 (383) | 76.4 (347) | 81.5 (753) | 75.3 (697) |
| A | 19.9 (92) | **25.6 (120)^b^** | 17.1 (79) | **23.6 (107)^b^** | 18.5 (171) | **24.7 (229)^c^** |
| Both sexes | *n =* 924 | *n =* 1000 | *n =* 1070 | *n =* 958 | *n =* 1994 | *n =* 1968 |
| T | 79.1 (731) | 76.2 (762) | 82.2 (880) | 76.4 (732) | 80.8 (1611) | 76.3 (1502) |
| A | 20.9 (193) | 23.8 (238) | 17.8 (190) | **23.6 (226)^d^** | 19.2 (383) | **23.7 (466)^c^** |
| Genotypes | **Genotypic frequency % (*n*)** | | | | | |
| Women | *n =* 231 | *n =* 268 | *n =* 304 | *n =* 253 | *n =* 535 | *n =* 521 |
| TT | 62.8 (145) | 60.4 (162) | 67.4 (205) | 59.3 (150) | 65.4 (350) | 60.1 (315) |
| AT | 30.7 (71) | 33.6 (90) | 28.6 (87) | 33.6 (85) | 29.5 (158) | 33.4 (175) |
| AA | 6.5 (15) | 5.2 (14) | 3.9 (12) | 6.7 (17) | 5 (27) | 5.9 (31) |
| AA/AT | 37.2 (86) | 39.1 (104) | 32.6 (99) | 40.5 (102) | 34.6 (185) | **39.5 (206)^a^** |
| Men | *n =* 232 | *n =* 235 | *n =* 231 | *n =* 227 | *n =* 462 | *n =* 463 |
| TT | 64.2 (149) | 54.9 (129) | 70.6 (163) | **57.3 (130)** | 67.4 (312) | **55.8 (259)** |
| AT | 31 (72) | 38.3 (90) | 24.7 (57) | **38.3 (87)** | 27.9 (129) | **38.6 (179)** |
| AA | 4.3 (10) | 6.4 (15) | 4.8 (11) | **4.4 (10)^c^** | 4.5 (21) | **5.4 (25)^c^** |
| AA/AT | 35.5 (82) | **44.9 (105)^b^** | 29.4 (68) | **42.7 (97)^c^** | 32.5 (150) | **44.1 (204)^d^** |
| Both sexes | *n =* 463 | *n =* 503 | *n =* 535 | *n =* 480 | *n =* 997 | *n =* 984 |
| TT | 63.5 (294) | 57.9 (291) | 68.8 (368) | **58.3 (280)** | 66.3 (662) | **58.1 (574)** |
| AT | 30.9 (143) | 35.8 (180) | 26.9 (144) | **35.8 (172)** | 28.8 (287) | **35.8 (354)** |
| AA | 5.4 (25) | 5.8 (29) | 4.3 (23) | **5.6 (27)^c^** | 4.8 (48) | **5.7 (56)^d^** |
| AA/AT | 36.4 (168) | **41.8 (209)^a^** | 31.2 (167) | **41.5 (199)^d^** | 33.6 (335) | **41.7 (410)^d^** |

Cases diagnosed with type 2 diabetes at ≤45 years were compared with controls aged ≤54 years and cases diagnosed with type 2 diabetes at ≥46 years were compared with controls aged ≥55 years.

Statistically significant values are highlighted in bold (Pearson chi square test).

^a^ *P* <0.1; ^b^ *P* <0.05, ^c^ *P* <0.01, ^d^ *P* <0.001.

**Table S5.** Association of SNP rs689 alleles (*n* = 3952)^a^ and genotypes (*n* = 1976)^b^ with T2D in the main case–control study stratified by age at T2D diagnosis and sex.

|  | Univariate logistic regression models | | | | | |
| --- | --- | --- | --- | --- | --- | --- |
|  | **T2D dx ≤45 years** | | **T2D dx ≥46 years** | | **All cases** | |
|  | **OR (95% CI)** | ***P* value** | **OR (95% CI)** | ***P* value** | **OR (95% CI)** | ***P* value** |
| rs689 alleles | | | | | | |
| Women | *n =* 462/532 | | *n =* 608/504 | | *n =* 1070/1042 | |
| T | 1 |  | 1 |  | 1 |  |
| A | 1.02 (0.75-1.38) | 0.90 | **1.38 (1.03-1.85)** | **0.029** | 1.19 (0.97-1.47) | 0.10 |
| Men | *n =* 462/468 | | *n =* 462/454 | | *n =* 924/926 | |
| T | 1 |  | 1 |  | 1 |  |
| A | **1.39 (1.02-1.89)** | **0.038** | **1.49 (1.08-2.07)** | **0.015** | **1.45 (1.16-1.81)** | **0.0012** |
| Both sexes | *n =* 924/1000 | | *n =* 1070/958 | | *n =* 1994/1968 | |
| T | 1 |  | 1 |  | 1 |  |
| A | 1.18 (0.95-1.47) | 0.13 | **1.43 (1.15-1.78)** | **0.0012** | **1.31 (1.12-1.52)** | **0.0006** |
| No. risk alleles^c^ | | | | | | |
| Women | *n =* 266 | | *n =* 252 | | *n =* 518 | |
| 0 | 1 |  | 1 |  | 1 |  |
| 1 | 1.13 (0.77-1.66) | 0.52 | 1.34 (0.93-1.92) | 0.12 | 1.23 (0.95-1.6) | 0.12 |
| 2 | 0.84 (0.39-1.79) | 0.64 | 1.94 (0.9-4.17) | 0.092 | 1.28 (0.74-2.18) | 0.37 |
| Men | *n =* 234 | | *n =* 227 | | *n =* 461 | |
| 0 | 1 |  | 1 |  | 1 |  |
| 1 | 1.44 (0.98-2.13) | 0.064 | **1.91 (1.28-2.87)** | **0.0017** | **1.67 (1.26-2.21)** | **0.0003** |
| 2 | 1.73 (0.75-3.99) | 0.20 | 1.14 (0.47-2.77) | 0.77 | 1.43 (0.78-2.62) | 0.24 |
| Both sexes | *n =* 500 | | *n =* 479 | | *n =* 979 | |
| 0 | 1 |  | 1 |  | 1 |  |
| 1 | 1.27 (0.97-1.67) | 0.084 | **1.57 (1.2-2.06)** | **0.0011** | **1.42 (1.17-1.72)** | **0.0003** |
| 2 | 1.17 (0.67-2.05) | 0.58 | 1.54 (0.87-2.75) | 0.14 | 1.35 (0.9-2.01) | 0.15 |

CI, confidence interval; dx, diagnosis; *n*, controls/cases; OR, odds ratio; T2D, type 2 diabetes.

Statistically significant values are highlighted in bold (Enter method, Wald test).

^a^ Included 1994 controls (chromosomes) (female, *n* = 1070; male, *n* = 924). Cases diagnosed at ≤45 years were compared with controls ≤54 years old (*n =* 924) and cases diagnosed at ≥46 years were compared with controls ≥55 years old (*n =* 1070).

^b^ Included 997 controls (female, *n* = 535; male, *n* = 462). Cases diagnosed at ≤45 years were compared with controls ≤54 years old (*n =* 462) and cases diagnosed at ≥46 years were compared with controls ≥55 years old (*n =* 535).

^c^ 0 = TT, 1 = AT, and 2 = AA.

**Table S6.** Frequency of rs689 alleles (*n =* 2376) and genotypes (*n =* 1188) in the replica case–control study stratified by age at type 2 diabetes diagnosis and sex.

|  | Controls ≤54 years | Cases dx ≤45 years | Controls ≥55 years | Cases dx ≥46 years | All controls | All cases |
| --- | --- | --- | --- | --- | --- | --- |
| Alleles | **Allelic frequency % (*n*)** | | | | | |
| Women | *n =* 486 | *n =* 296 | *n =* 360 | *n =* 294 | *n =* 846 | *n =* 594 |
| T | 77.6 (377) | 77.4 (229) | 76.9 (277) | 75.5 (222) | 77.3 (654) | 76.6 (455) |
| A | 22.4 (109) | 22.6 (67) | 23.1 (83) | 24.5 (72) | 22.7 (192) | 23.4 (139) |
| Men | *n =* 158 | *n =* 330 | *n =* 186 | *n =* 260 | *n =* 344 | *n =* 592 |
| T | 80.4 (127) | 73.3 (242) | 76.3 (142) | 77.3 (201) | 78.2 (269) | 75 (444) |
| A | **19.6 (31)** | **26.7 (88)^a^** | 23.7 (44) | 22.7 (59) | 21.8 (75) | 25.0 (148) |
| Both sexes | *n =* 644 | *n =* 626 | *n =* 546 | *n =* 554 | *n =* 1190 | *n =* 1186 |
| T | 78.3 (504) | 75.2 (471) | 76.7 (419) | 76.4 (423) | 77.6 (923) | 75.8 (899) |
| A | 21.7 (140) | 24.8 (155) | 23.3 (127) | 23.6 (131) | 22.4 (267) | 24.2 (287) |
| Genotypes | **Genotypic frequency % (*n*)** | | | | | |
| Women | *n =* 243 | *n =* 144 | *n =* 180 | *n =* 153 | *n =* 423 | *n =* 297 |
| TT | 61.3 (149) | 63.9 (92) | **56.7 (102)** | **59.5 (91)** | **59.3 (251)** | **61.6 (183)** |
| AT | 32.5 (79) | 28.5 (41) | **40.6 (73)** | **31.4 (48)** | **35.9 (152)** | **30.0 (89)** |
| AA | 6.2 (15) | 7.6 (11) | **2.8 (5)** | **9.2 (14)^b^** | **4.7 (20)** | **8.4 (25)^a^** |
| AA/AT | 38.7 (94) | 36.1 (52) | 43.3 (78) | 40.5 (62) | 40.7 (172) | 38.4 (114) |
| Men | *n =* 79 | *n =* 164 | *n =* 93 | *n =* 132 | *n =* 172 | *n =* 296 |
| TT | 65.8 (52) | 52.4 (86) | 54.8 (51) | 58.3 (77) | 59.9 (103) | 55.1 (163) |
| AT | 29.1 (23) | 42.1 (69) | 43.0 (40) | 37.1 (49) | 36.6 (63) | 39.9 (118) |
| AA | 5.1 (4) | 5.5 (9) | 2.2 (2) | 4.5 (6) | 3.5 (6) | 5.1 (15) |
| AA/AT | **34.2 (27)** | **47.6 (78)^b^** | 45.2 (42) | 41.7 (55) | 40.1 (69) | 44.9 (133) |
| Both sexes | *n =* 322 | *n =* 308 | *n =* 273 | *n =* 285 | *n =* 595 | *n =* 593 |
| TT | 62.4 (201) | 57.8 (178) | **56.0 (153)** | **58.9 (168)** | 59.5 (354) | 58.3 (346) |
| AT | 31.7 (102) | 35.7 (110) | **41.4 (113)** | **34 (97)** | 36.1 (215) | 34.9 (207) |
| AA | 5.9 (19) | 6.5 (20) | **2.6 (7)** | **7.0 (20)^b^** | 4.4 (26) | 6.7 (40) |
| AA/AT | 37.6 (121) | 42.2 (130) | 44.0 (120) | 41.1 (117) | 40.5 (241) | 41.7 (247) |

Statistically significant values are highlighted in bold (Pearson chi square test).

^a^ *P* <0.1; ^b^ *P* <0.05, ^c^ *P* <0.01.

**Table S7.** Association of SNP rs689 alleles and genotypes with type 2 diabetes in the replica case–control study stratified by age at type 2 diabetes diagnosis and sex.

| Groups | Alleles/ Genotypes | Univariate logistic regression models | | | | | |
| --- | --- | --- | --- | --- | --- | --- | --- |
|  |  | **Dx ≤45 years** | | **Dx ≥46 years** | | **All cases** | |
|  |  | **OR (95% CI)** | ***P* value** | **OR (95% CI)** | ***P* value** | **OR (95% CI)** | ***P* value** |
| Alleles (*n =* 2376) | | | | | | | |
| Women |  | *n =* 486/296 | | *n =* 360/294 | | *n =* 846/594 | |
|  | T | 1 |  | 1 |  | 1 |  |
|  | A | 1.012 (0.72-1.43) | 0.946 | 1.082 (0.75-1.55) | 0.668 | 1.196 (0.87-1.64) | 0.269 |
| Men |  | *n =* 158/330 | | *n =* 186/260 | | *n =* 344/592 | |
|  | T | 1 |  | 1 |  | 1 |  |
|  | A | **1.49 (0.94-2.37)** | **0.09** | 0.947 (0.61-1.48) | 0.812 | 1.041 (0.81-1.33) | 0.754 |
| Both sexes |  | *n =* 644/626 | | *n =* 546/554 | | *n =* 1190/1186 | |
|  | T | 1 |  | 1 |  | 1 |  |
|  | A | 1.185 (0.91-1.54) | 0.203 | 1.022 (0.77-1.35) | 0.88 | 1.104 (0.91-1.33) | 0.310 |
| Genotypes (*n =* 1188) | | | | | | | |
| Women |  | *n* = 243/144 | | *n* = 180/153 | | *n* = 423/297 | |
|  | TT | 1 |  | 1 |  | 1 |  |
|  | AA/AT | 0.896 (0.58-1.37) | 0.614 | 0.891 (0.58-1.38) | 0.605 | 0.909 (0.67-1.23) | 0.539 |
| Men |  | *n* = 79/164 | | *n* = 93/132 | | *n* = 172/296 | |
|  | TT | 1 |  | 1 |  | 1 |  |
|  | AA/AT | **1.747 (1-3.05)** | **0.049** | 0.867 (0.51-1.48) | 0.602 | 1.218 (0.83-1.78) | 0.311 |
| Both sexes |  | *n* = 322/308 | | *n* = 273/285 | | *n* = 595/593 | |
|  | TT | 1 |  | 1 |  | 1 |  |
|  | AA/AT | 1.213 (0.88-1.67) | 0.236 | 0.888 (0.63-1.24) | 0.488 | 1.049 (0.83-1.32) | 0.687 |

CI, confidence interval; Dx, diagnosis; *n* = controls/cases; OR, odds ratio; SNP, small nucleotide polymorphism*.*

Statistically significant values are highlighted in bold.

**Table S8.** Frequency of rs689 alleles (*n* = 2344) and genotypes (*n* = 1172) in the cross-sectional study stratified by age and sex.

|  | Controls ≤47 years | Cases dx ≤47 years | Controls ≥48 years | Cases dx ≥48 years | All controls | All cases |
| --- | --- | --- | --- | --- | --- | --- |
| Alleles | **Allelic frequency % (*n*)** | | | | | |
| Women | *n* = 778 | *n* = 62 | *n* = 800 | *n* = 82 | *n* = 1578 | *n* = 144 |
| T | 79.7 (620) | 79.0 (49) | 81.9 (655) | 82.9 (68) | 80.8 (1275) | 81.3 (117) |
| A | 20.3 (158) | 21.0 (13) | 18.1 (145) | 17.1 (14) | 19.2 (303) | 18.8 (27) |
| Men | *n* = 290 | *n* = 34 | *n* = 268 | *n* = 30 | *n* = 558 | *n* = 64 |
| T | 83.8 (243) | 64.7 (22) | 76.1 (204) | 76.7 (23) | 80.1 (447) | 70.3 (45) |
| A | 16.2 (47) | **35.3 (12)^c^** | 23.9 (64) | 23.3 (7) | 19.9 (111) | **29.7 (19)^a^** |
| Both sexes | *n* = 1068 | *n* = 96 | *n* = 1068 | *n* = 112 | *n* = 2136 | *n* = 208 |
| T | 80.8 (863) | 74.0 (71) | 80.4 (859) | 81.3 (91) | 80.6 (1722) | 77.9 (162) |
| A | 19.2 (205) | 26.0 (25) | 19.6 (209) | 18.8 (21) | 19.4 (414) | 22.1 (46) |
| Genotypes | **Genotypic frequency % (*n*)** | | | | | |
| Women | *n* = 389 | *n* = 31 | *n* = 400 | *n* = 41 | *n* = 789 | *n* = 72 |
| TT | 63.2 (246) | 64.5 (20) | 66.3 (265) | 70.7 (29) | 64.8 (511) | 68.1 (49) |
| AT | 32.9 (128) | 29.0 (9) | 31.3 (125) | 24.4 (10) | 32.1 (253) | 26.4 (19) |
| AA | 3.9 (15) | 6.5 (2) | 2.5 (10) | 4.9 (2) | 3.2 (25) | 5.6 (4) |
| Men | *n* = 145 | *n* = 17 | *n* = 134 | *n* = 15 | *n* = 279 | *n* = 32 |
| TT | 68.3 (99) | **41.2 (7)** | 58.2 (78) | 66.7 (10) | 63.4 (177) | **53.1 (17)** |
| AT | 31.0 (45) | **47.1 (8)** | 35.8 (48) | 20.0 (3) | 33.3 (93) | **34.4 (11)** |
| AA | 0.7 (1) | **11.8 (2)^c^** | 6.0 (8) | 13.3 (2) | 3.2 (9) | **12.5 (4)^b^** |
| Both sexes | *n* = 534 | *n* = 48 | *n* = 534 | *n* = 56 | *n* = 1068 | *n* = 104 |
| TT | 64.6 (345) | 56.3 (27) | 64.2 (343) | 69.6 (39) | 64.4 (688) | **63.5 (66)** |
| AT | 32.4 (173) | 35.4 (17) | 32.4 (173) | 23.2 (13) | 32.4 (346) | **28.8 (30)** |
| AA | 3.0 (16) | 8.3 (4) | 3.4 (18) | 7.1 (4) | 3.2 (34) | **7.7 (8)^a^** |

Dx, diagnosis.

Statistically significant values are highlighted in bold (Pearson chi square test).

^a^ *P* <0.1; ^b^ *P* <0.05, ^c^ *P* <0.01.

**Table S9.** Association of SNP rs689 alleles (*n* = 2344) and genotypes (*n* = 1172) with type 2 diabetes in the cross-sectional study stratified by age and sex.

| Groups | No. risk alleles | Univariate logistic regression models | | | | | |
| --- | --- | --- | --- | --- | --- | --- | --- |
|  |  | **Dx ≤47 years** | | **Dx ≥48 years** | | **All cases** | |
|  |  | **OR (95% CI)** | ***P* value** | **OR (95% CI)** | ***P* value** | **OR (95% CI)** | ***P* value** |
| Alleles | | | | | | | |
| Women |  | *n* = 778/62 | | *n* = 800/82 | | *n* = 1578/144 | |
|  | 0 | 1 |  | 1 |  | 1 |  |
|  | 1 | 1.04 (0.55-1.97) | 0.90 | 0.93 (0.51-1.7) | 0.81 | 0.97 (0.63-1.5) | 0.90 |
| Men |  | *n* = 290/34 | | *n* = 268/30 | | *n* = 558/64 | |
|  | 0 | 1 |  | 1 |  | 1 |  |
|  | 1 | **2.82 (1.31-6.09)** | **0.008** | 0.97 (0.4-2.37) | 0.95 | **1.7 (0.96-3.02)** | **0.07** |
| Both sexes |  | *n* = 1068/96 | | *n* = 1068/112 | | *n* = 2136/208 | |
|  | 0 | 1 |  | 1 |  | 1 |  |
|  | 1 | 1.48 (0.92-2.4) | 0.11 | 0.95 (0.58-1.56) | 0.84 | 1.18 (0.84-1.67) | 0.34 |
| Genotypes | | | | | | | |
| Women |  | *n* = 389/31 | | *n* = 400/41 | | *n* = 789/72 | |
|  | 0 | 1 |  | 1 |  | 1 |  |
|  | 1 | 0.86 (0.38-1.95) | 0.73 | 0.73 (0.35-1.55) | 0.41 | 0.78 (0.45-1.36) | 0.38 |
|  | 2 | 1.64 (0.35-7.68) | 0.53 | 1.83 (0.38-8.75) | 0.45 | 1.67 (0.56-4.99) | 0.36 |
| Men |  | *n* = 145/17 | | *n* = 134/15 | | *n* = 279/32 | |
|  | 0 | 1 |  | 1 |  | 1 |  |
|  | 1 | **2.51 (0.86-7.36)** | **0.092** | 0.49 (0.13-1.86) | 0.29 | 1.23 (0.55-2.74) | 0.61 |
|  | 2 | **28.29 (2.28-351.51)** | **0.0093** | 1.95 (0.36-10.5) | 0.44 | **4.63 (1.29-16.62)** | **0.019** |
| Both sexes |  | *n* = 534/48 | | *n* = 534/56 | | *n* = 1068/104 | |
|  | 0 | 1 |  | 1 |  | 1 |  |
|  | 1 | 1.26 (0.67-2.37) | 0.48 | 0.66 (0.34-1.27) | 0.21 | 0.9 (0.58-1.42) | 0.66 |
|  | 2 | **3.19 (1-10.23)** | **0.05** | 1.95 (0.63-6.07) | 0.25 | **2.45 (1.09-5.52)** | **0.03** |

CI, confidence interval; Dx, diagnosis; *n* = controls/cases; OR, odds ratio; SNP, small nucleotide polymorphism*.*

Statistically significant values are highlighted in bold.

**Table S10.** Association of *TH01*/rs689 haplotypes with type 2 diabetes stratified by age at type 2 diabetes diagnosis and sex (*n* = 3952)^a^.

| *TH01*/rs689 haplotypes | Univariate logistic regression models | | | | | |
| --- | --- | --- | --- | --- | --- | --- |
|  | **Dx ≤45 years** | | **Dx ≥46 years** | | **All cases** | |
|  | **OR (95% CI)** | ***P* value** | **OR (95% CI)** | ***P* value** | **OR (95% CI)** | ***P* value** |
| Women | *n* = 532 | | *n* = 504 | | *n* = 1036 | |
| ≤7R /T or A | 1 |  | 1 |  | 1 |  |
| ≥8R /T | **0.53 (0.36-0.77)** | **0.0011** | 1.51 (1-2.29) | 0.052 | 0.9 (0.68-1.19) | 0.45 |
| ≥8R /A | 0.96 (0.7-1.33) | 0.82 | **1.45 (1.07-1.98)** | **0.018** | 1.2 (0.96-1.5) | 0.10 |
| Men | *n* = 468 | | *n* = 454 | | *n* = 922 | |
| ≤7R /T or A | 1 |  | 1 |  | 1 |  |
| ≥8R /T | **1.68 (1.05-2.69)** | **0.031** | **1.85 (1.19-2.87)** | **0.0061** | **1.76 (1.28-2.42)** | **0.0006** |
| ≥8R /A | **1.37 (0.98-1.9)** | **0.063** | **1.44 (1.02-2.02)** | **0.037** | **1.41 (1.11-1.79)** | **0.0043** |
| Both sexes | *n* = 1000 | | *n* = 958 | | *n* = 1958 | |
| ≤7R /T or A | 1 |  | 1 | 0 | 1 |  |
| ≥8R /T | 0.86 (0.64-1.15) | 0.30 | **1.67 (1.24-2.26)** | **0.0008** | 1.2 (0.98-1.48) | 0.082 |
| ≥8R /A | 1.15 (0.91-1.44) | 0.24 | **1.45 (1.15-1.82)** | **0.0016** | **1.3 (1.1-1.52)** | **0.0016** |

CI, confidence interval; Dx, diagnosis; OR, odds ratio; R, repeats*.*

^a^ A total of 1994 controls (chromosomes) were included (female, *n* = 1070; male, *n* = 924). Cases diagnosed at ≤45 years old were compared with controls ≤54 years old (*n =* 924) and cases diagnosed at ≥46 years old were compared with controls ≥55 years old (*n =* 1070).

Statistically significant values are highlighted in bold (Enter method, Wald test).

**Table S11.** Fasting plasma insulin levels in the main case–control study stratified by age at type 2 diabetes diagnosis, sex and *TH01* and rs689 alleles (*n* = 2564).

| **Locus** | **Alleles** | **Fasting insulin (mIU/mL)** | | | | | | | | | | | |
| --- | --- | --- | --- | --- | --- | --- | --- | --- | --- | --- | --- | --- | --- |
|  |  | **Controls ≤54 years** | | **Cases Dx ≤45 years** | | **Controls ≥55 years** | | **Cases Dx ≥46 years** | | **All controls** | | **All cases** | |
|  |  | ***n*** | **Median (IQR)** | ***n*** | **Median**  **(IQR)** | ***n*** | **Median (IQR)** | ***n*** | **Median**  **(IQR)** | ***n*** | **Median (IQR)** | ***n*** | **Median**  **(IQR)** |
| **Women** | | | | | | | | | | | | | |
| ***TH01*** | ≤7R | 269 | 7.4 (5-10.5) | 310 | **10.7 (6.5-18)^c^** | 219 | 6.6 (4.3-9.5) | 346 | **8.9 (5.3-14.4)^c^** | 488 | 6.9 (4.8-9.8) | 660 | 9.5 (5.7-16.2) |
|  | ≥8R | 153 | 6.8 (4.9-11) | 140 | **12.4 (7.2-19.7)^a^** | 71 | 6.3 (5-10.7) | 158 | **10.1 (5.6-16.5)^a^** | 224 | 6.7 (4.9-10.8) | 300 | 10.9 (6.3-19) |
|  | Total | 422 | 7.1 (5-10.6) | 450 | **11 (6.7-18.1)^d^** | 290 | 6.4 (4.3-9.5) | 504 | **9 (5.4-15.3)^d^** | 712 | 6.8 (4.8-10) | 960 | 10.2 (5.8-17) |
| **rs689** | T | 329 | 7.3 (5-10.6) | 343 | **10.8 (6.7-18)^c^** | 241 | 6.3 (4.3-9.5) | 383 | **9 (5.3-15.2)^c^** | 570 | 6.8 (4.7-9.9) | 732 | 10 (5.8-16.5) |
|  | A | 93 | 6.5 (4.9-10.3) | 105 | 11.7 (6.6-19.8) | 49 | 7.2 (5.1-11.2) | 119 | 10.2 (5.4-15.5) | 142 | 6.9 (5-10.7) | 224 | 10.7 (5.9-17.9) |
| **Men** | | | | | | | | | | | | | |
| ***TH01*** | ≤7R | 190 | 6.4 (5-10) | 176 | **10 (5.7-18.2)^d^** | 100 | 6.2 (4.4-8.7) | 148 | **6.8 (3.7-10.4)^d^** | 290 | **6.3 (4.7-9.5)** | 325 | 8.5 (4.4-14.5) |
|  | ≥8R | 72 | 7 (5.2-10.1) | 82 | **10.7 (6.5-19)^c^** | 40 | 8 (4.2-13.7) | 82 | **8.2 (4.5-12.3)^c^** | 112 | **7.4 (4.9-11.3)** | 165 | 9.1 (5.3-14) |
|  | Total | 262 | 6.9 (5-10) | 258 | **10.1 (6-18.2)^d^** | 140 | 6.5 (4.2-10.1) | 230 | **7.1 (3.8-10.7)^d^** | 402 | 6.7 (4.7-10) | 490 | 8.7 (4.8-14) |
| **rs689** | T | 206 | 6.8 (5.1-9.9) | 187 | **10 (6-17.8)^d^** | 117 | 6.3 (4.5-9.4) | 170 | **6.9 (3.8-10.4)^d^** | 323 | 6.5 (4.7-9.9) | 358 | 8.6 (4.5-13.3) |
|  | A | 56 | 7 (4.6-10.3) | 69 | **10.3 (6.3-19)^b^** | 23 | 8.6 (4.2-14) | 60 | **8.2 (3.8-13.2)^b^** | 79 | 7.2 (4.5-12.5) | 130 | 9.4 (5.1-16) |
| **Both sexes** | | | | | | | | | | | | | |
| ***TH01*** | ≤7R | 459 | 7 (5-10.2) | 486 | **10.5 (6.2-18)^d^** | 319 | 6.4 (4.3-9.5) | 494 | **8.1 (4.5-13.9)^d^** | 778 | 6.7 (4.7-9.8) | 985 | 9.2 (5.3-15.9) |
|  | ≥8R | 225 | 7 (4.9-10.3) | 222 | **12.1 (6.8-19.6)^c^** | 111 | 6.8 (4.6-11.8) | 240 | **9.1 (5.2-15)^c^** | 336 | 7 (4.9-10.8) | 465 | 10.3 (5.8-17.1) |
| **rs689** | T | 535 | 7 (5-10.4) | 530 | **10.8 (6.5-18)^c^** | 358 | 6.3 (4.3-9.5) | 553 | **8.5 (4.7-13.9)^c^** | 893 | 6.7 (4.7-9.9) | 1090 | 9.4 (5.4-16) |
|  | A | 149 | 6.9 (4.8-10.3) | 174 | 11.4 (6.5-19.6) | 72 | 7.4 (4.9-12) | 179 | 9.4 (5-15) | 221 | 7 (4.8-10.7) | 354 | 10.3 (5.6-17.1) |
| **Total** |  | 684 | 7 (5-10.3) | 708 | **10.8 (6.5-18.2)^d^** | 430 | 6.4 (4.3-9.9) | 734 | **8.5 (4.8-14.1)^d^** | 1114 | 6.8 (4.8-10) | 1450 | **9.5 (5.5-16.2)^d^** |

Dx, diagnosis; IQR, interquartile range.

Cases diagnosed at ≤45 years old were compared with controls ≤54 years old and cases diagnosed at ≥46 years old were compared with controls ≥55 years old. Statistically significant values are highlighted in bold (Pearson-chi square).

^a^ *P* <0.05; ^b^ *P* <0.01; ^c^ *P* <0.001; ^d^ *P* <0.0001.
